# Novel Genetic Risk Loci for Pancreatic Ductal Adenocarcinoma Identified in a Genome-wide Study of African Ancestry Individuals

**DOI:** 10.64898/2026.04.21.26351329

**Authors:** Candelaria Vergara, Zhanmo Ni, Jun Zhong, David McKean, Katelyn E. Connelly, Samuel O. Antwi, Alan A Arslan, Paige M. Bracci, Mengmeng Du, Steven Gallinger, Jeanine Genkinger, Christopher A Haiman, Manal Hassan, Rayjean J. Hung, Chad Huff, Charles Kooperberg, Fay Kastrinos, Loic LeMarchand, WooHyung Lee, Shannon M. Lynch, Stephen C Moore, Ann L. Oberg, Margaret A Park, Jennifer B Permuth, Harvey A. Risch, Paul Scheet, Ann Schwartz, Xiao-Ou Shu, Rachael Z Stolzenberg-Solomon, Brian M Wolpin, Wei Zheng, Demetrius Albanes, Gabriella Andreotti, William R. Bamlet, Laura Beane-Freeman, Sonja I Berndt, Paul Brennan, Julie E Buring, Natalia Cabrera-Castro, Daniele Campa, Federico Canzian, Stephen J Chanock, Yu Chen, Charles C Chung, A. Heather Eliassen, J. Michael Gaziano, Graham G Giles, Edward L Giovannucci, Michael Goggins, Phyllis J Goodman, Belynda Hicks, Amy Hutchinson, Miranda R Jones, Verena Katzke, Manolis Kogevinas, Robert C. Kurtz, Daniel Laheru, I-Min Lee, Núria Malats, Roger Milne, Lorelei Mucci, Rachel E. Neale, Irene Orlow, Alpa V Patel, Laia Peruchet, Ulrike Peters, Miquel Porta, Kari G. Rabe, Francisco X Real, Fulvio Ricceri, Nathaniel Rothman, Howard D Sesso, Veronica W Setiawan, Debra Silverman, Melissa C Southey, Meir J Stampfer, Geoffrey S Tobias, Caroline Um, Kala Visvanathan, Jean Wactawski-Wende, Nicolas Wentzensen, Walter C Willett, Herbert Yu, Peter Kraft, Priya Duggal, Laufey T Amundadottir, Alison P. Klein

## Abstract

Pancreatic cancer disproportionately affects Black individuals in the United States, but they have limited representation in genetic studies of pancreatic ductal adenocarcinoma (PDAC). To address this gap, we performed admixture mapping and genome-wide association analysis (GWAS) in genetically inferred African ancestry individuals (1,030 cases and 889 controls). Admixture mapping identified three regions with a significantly higher proportion of African ancestry in cases compared to controls (5q33.3, 10p1, 22q12.3). GWAS identified a genome-wide significant association at 5p15.33 (*CLPTM1L*, rs383009:T>C, T Allele Frequency=0.51, OR:1.45, P value=1.24×10^-8^), a locus previously associated with PDAC. Known loci at 5p15.33, 7q32.3, 8q24.21 and 7q25.1 also replicated (P value <0.01). Multi-ancestral fine-mapping identified two potential causal SNPs (rs3830069 and rs2735940) at 5p15.33. Collectively these findings identified novel PDAC risk loci and expanded our understanding of this deadly cancer in underrepresented populations, emphasizing the multifactorial nature of PDAC risk including inherited genetic and non-genetic factors.

**Statement of Significance:** To understand how genetic variation contributes to PDAC risk in Black people in North American, we studied individuals of genetically-inferred African ancestry. We identified novel risk loci and differences in the contribution of known loci. This demonstrates that ancestry-informed genetic analyses improve our understanding of PDAC risk and enhances discovery.

## Introduction

Pancreatic cancer is currently the third leading cause of cancer death in the United States [1]. An estimated 67,530 new pancreatic cancer cases are expected to be diagnosed in the United States in 2026 [1]. Globally, age-adjusted incidence rates of pancreatic cancer have increased over the past two decades from 5 per 100,000 to 5.7 per 100,000 [2]. In the United States, the age-adjusted incidence rates of pancreatic cancer among Black individuals are higher than among White individuals with rate ratios of 1.14 and 1.13 for men and women respectively, although differences in rates between Black and White individuals have decreased in recent years [2, 3]. While some studies indicate that observed variation in age-adjusted incidence rates by race and ethnicity are due to different prevalence of key non-genetic risk factors (i.e., smoking, diabetes, heavy alcohol intake, high BMI), recent multiethnic cohort studies have indicated that the variation in incidence rates of pancreatic cancer may not be fully explained by differences in the distribution of those risk factors [4, 5]. Differences in genetic factors have not been well studied.

Genome-wide association studies have demonstrated that common genetic variation contributes to pancreatic ductal adenocarcinoma (PDAC), the most common pancreatic cancer sub-type. In populations of European ancestry, 22 genomic regions are associated with PDAC, including 1q32.1, 1p36.33, 2p13.3, 3q29, 5p15.33, 7p14.1, 8q21.11, 8q24.21, 9q34.2, 13q12.2, 13q22.1, 16q23.1,17q12, 17q25.1,18q21.32, and 22q12.1 [6–10] and yet these regions only explain 4.1% of the PDAC liability [11], suggesting more genetic loci are involved. Additional risk regions have been identified in Japanese populations including 6p25.3, 12p11.21, 7q36.2, 13q12.2, 13q221, and 16p12.3 [12]; and in Chinese populations 21q21.3, 5p13.1, 21q22.3, 22q13.32, and 10q26.11 [13]. However, large-scale studies of PDAC risk in North American Black individuals who represent both African and European ancestry populations remain limited.

To address this gap, we investigated the genetic architecture of PDAC in individuals with inferred African ancestry, with the goal of identifying novel PDAC risk loci. Using whole genome sequencing data from 1,030 PDAC cases and 889 controls, we performed genome-wide admixture mapping to assess local ancestry associations with pancreatic cancer status and performed a SNP-based GWAS to identify additional risk loci. We also evaluated genomic regions previously reported as PDAC-associated loci in European ancestry populations using multi-ancestral fine-mapping to refine putative causal variants within these regions.

## Methods

### Study Population

Cases and controls, who self-reported as Black/African American were participants of the Pancreatic Cancer Case Control and Pancreatic Cancer Cohort Consortia as well as controls from the INHALE Study as described in Supplementary Materials and Methods and in **Supplemental Table 1**. Written informed consent was obtained from participants in each study, and the study protocol was approved by the IRB at Johns Hopkins School of Medicine and the National Cancer Institute, the National Institutes of Health.

Cases were individuals with a diagnosis of PDAC as defined by ICD-O codes (C25.0-25.9, excluding 25.4) and/or examination of diagnostic specimens, pathology reports, medical records, and/or death certificates. Controls were individuals without a diagnosis of PDAC and when possible, frequency-matched by age to the cases and geographic region. Phenotype data from the different studies were harmonized at Johns Hopkins before analyses.

Participants were included in the genetic analyses based on inferred African ancestry using genome-wide genotype data. Only individuals with principal components clustering with African-ancestry genetic profiles derived from 1000 Genomes, were retained to provide relative homogeneity in genetic architecture for the identification of PDAC-associated loci. In addition, for fine-mapping we included summary statistics data from our prior GWAS studies, PanScan I-III and PanC4 GWAS studies of European genetic ancestry individuals in the same cohorts [6–10].

### Quality control and Ancestry Estimation

Germline 30X whole genome sequencing was conducted at the Center for Inherited Disease Research. Alignment, variant call, and sequencing quality control are described in Supplementary Material. Standard genome-wide quality control was performed for the samples and markers included in the GWAS analysis [14] as described in detail in the Supplementary Material. In brief, we excluded markers with a minor allele frequency (MAF < 0.01), genotyping rate < 95% and/or deviations from Hardy Weinberg equilibrium (P value <1×10^-9^). In total,19,898,673 markers were included in the GWAS analysis. For admixture mapping, we used the same standard quality control, in this case excluding markers with a MAF < 0.05 as well as markers that were not present in the 1000 Genomes phase 3 reference populations. In total, we used 7,663,144 autosomal markers to estimate global and local ancestry. The Genome Reference Consortium Human 38 (GRCh38) was used as reference.

After calling the variants in 1,968 unique experimental samples, we excluded samples with genotyping rate <95%, sex discrepancies, and first- and second-degree relatedness as described. We performed principal component analysis to estimate and adjust the association analyses results based on population structure using the algorithm implemented in EIGENSOFT [15, 16] (See Supplementary Material and Methods for details). The first component captured 22.14% of the variance and was sufficient to differentiate samples from African- and European-ancestry individuals (**Supplemental Figure 1**).

To estimate global and local genome-wide ancestry of the participants, we used a two-way admixture model incorporating African (YRI) and European (CEU) reference individuals from 1000 Genomes Phase 3 Reference Panel [17, 18]. Local ancestry was estimated genome-wide using random-forest discriminative machine-learning methods, combined with a conditional random field model of the linear chromosome as implemented in RFMIx [17]. To ensure consistency in the admixture and GWAS analyses we limited to individuals who clustered with the YRI or who had a global African ancestry greater than 20% (n=35 excluded). Details are available in Supplementary Materials and Methods.

### Statistical Analysis

#### Demographic variables

Demographic variables such as categories of age and sex were compared between cases and controls using chi-square tests. A student t-test was used to compare global ancestry averages between cases and controls. A two-sided nominal P value < 0.05 was considered statistically significant.

#### Association of Local Ancestry with PDAC

Association testing between PDAC and local African ancestry was conducted via admixture mapping using logistic mixed model implemented in LLAMA [18] within the GENESIS (GENetic EStimation and Inference in Structured samples) R package [19], which provides methodology for estimating, inferring, and accounting for population and pedigree structure by including principal components and a genetic relatedness matrix (GRM) (see Supplementary Methods). Each local-ancestry segment was encoded as 0, 1, or 2 based on the number of inferred African haplotypes. Models were adjusted for age, sex, and global inferred African ancestry as fixed effects, with the genetic relatedness matrix included as a random effect (as described in **Table 1)** [19]. To account for multiple testing, we calculated the genome wide significance threshold for admixture mapping using the Significance Threshold Estimation for Admixture Mapping (STEAM) algorithm [20] using GRCh38 genetic maps, with 1,085,901 genomic segments and assuming 6 generations since the event of admixture, resulting in a significance cutoff of two-sided P value < 1.27 x10^-5^.

**Table 1.**
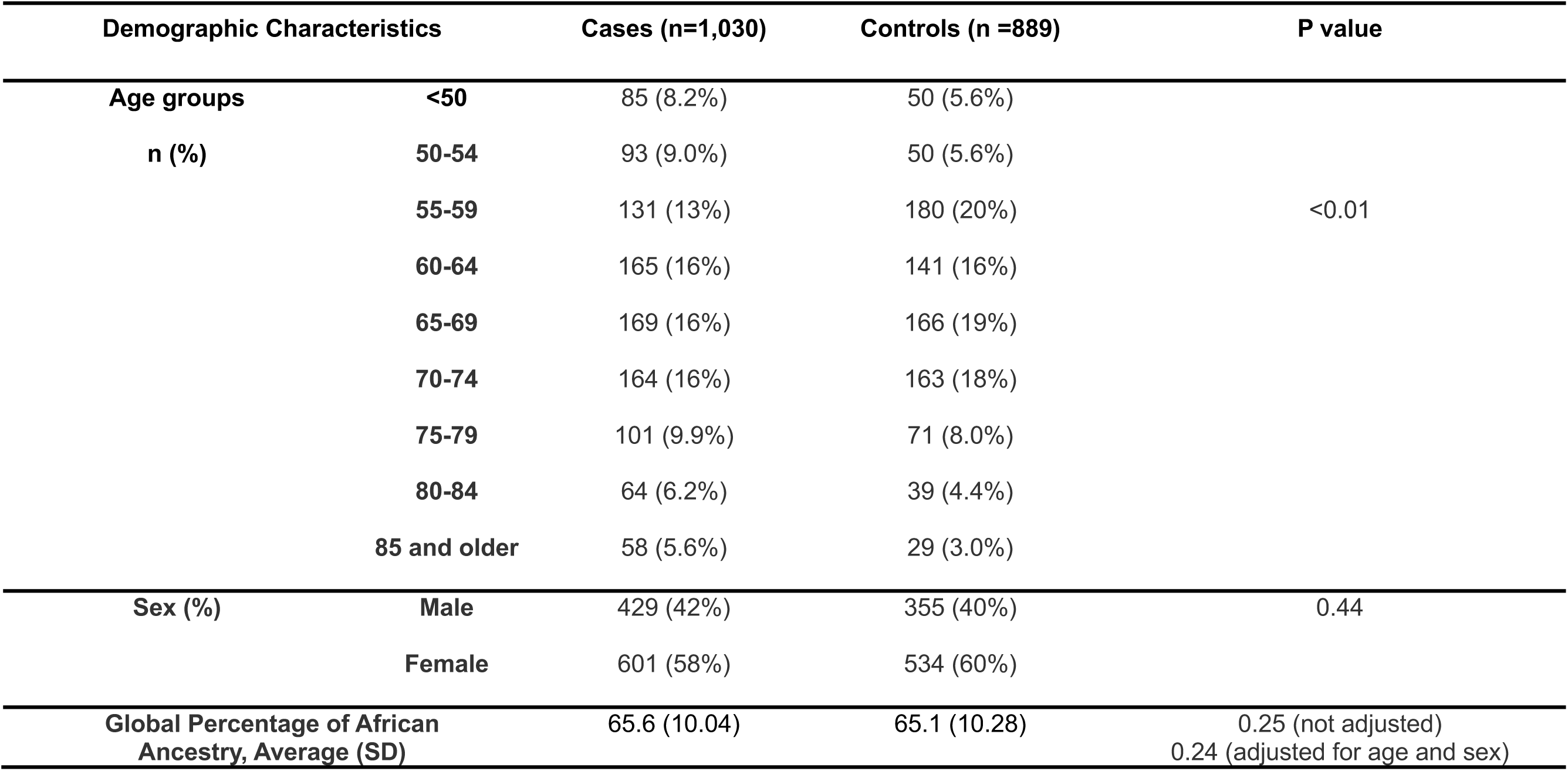
Demographic characteristics of the study population by case-control status.

| Demographic Characteristics |  | Cases (n=1,030) | Controls (n =889) | P value |
| --- | --- | --- | --- | --- |
| Age groups<br>n (%) | <50 | 85 (8.2%) | 50 (5.6%) | <0.01 |
|  | 50-54 | 93 (9.0%) | 50 (5.6%) |  |
|  | 55-59 | 131 (13%) | 180 (20%) |  |
|  | 60-64 | 165 (16%) | 141 (16%) |  |
|  | 65-69 | 169 (16%) | 166 (19%) |  |
|  | 70-74 | 164 (16%) | 163 (18%) |  |
|  | 75-79 | 101 (9.9%) | 71 (8.0%) |  |
|  | 80-84 | 64 (6.2%) | 39 (4.4%) |  |
|  | 85 and older | 58 (5.6%) | 29 (3.0%) |  |
| Sex (%) | Male | 429 (42%) | 355 (40%) | 0.44 |
|  | Female | 601 (58%) | 534 (60%) |  |
| Global Percentage of African Ancestry, Average (SD) |  | 65.6 (10.04) | 65.1 (10.28) | 0.25 (not adjusted)<br>0.24 (adjusted for age and sex) |

#### Genome Wide SNP Association Analysis

Association between SNPs and PDAC risk was tested using a generalized mixed model implemented in the program SAIGE (Scalable and Accurate Implementation of GEneralized mixed model) [21] including a GRM-based on the raw genotypes, age in 5-year age categories (as described in **Table 1**), sex, and one principal component (see Supplementary Materials and Methods). Genetic variant associations with two-sided P value < 5 x 10^-8^ were considered genome-wide significant and variants with two-sided P value < 1 x 10^-7^ were considered suggestive. Variants with prior genome-wide significant evidence of association were considered replicated if their two-sided P values were less than 0.01.

#### Multi-Ancestry Fine-Mapping

Given that genome-wide association studies often identify many associated variants in a given region due the underlying correlation (linkage disequilibrium) between markers, further fine-mapping is needed to narrow down the potentially causal variants from correlated non-causal variants. Leveraging the different LD patterns across ancestries can improve fine-mapping. To further narrow sets of putative causal variants in associated regions, we conducted fine-mapping using the algorithm implemented in the multi-ancestry sum of the single effects models program (MESuSiE) [22]. MESuSie leverages both shared ancestry as well as ancestry specific LD patterns to obtain posterior inclusion probabilities (PIP) that a given variant is “causal“. A PIP > 0.5 was used as threshold to identify potential causal variants [22]. We performed fine-mapping analyses using our GWAS results from individuals of African ancestry together with summary statistics from our prior GWAS in individuals of European ancestry [6]. The European-ancestry data included genotyping from PanScan I-III and PanC4 studies [6] For the European GWAS dataset, genotype imputation was conducted using the TOPMed reference panel [23] (version R2 on human genome build GRCh38) via the TOPMed Imputation Server [24] and association analyses were performed under a multivariable logistic regression model using the additive approach in SNPTEST (v2.5.4-beta3) [25] adjusting for age, sex, global ancestry, and study phase. Additional methodologic details are provided in the Supplementary Materials.

We fine-mapped genomic regions with genome-wide significant or marginal association signals in any of the groups **(Supplemental Table 2)** and loci identified in our admixture mapping analyses (chr5q33.3, ch22q12.3, chr10p11) **(Table 2).** A posterior inclusion probability (PIP) exceeding the 0.5 threshold was determined as putative causal and further interrogated in Genotype-Tissue Expression (GTEx) project database [26], v10 Release via the GTEx portal (https://gtexportal.org/home/) for potential regulatory or functional effects.

**Table 2.** Results of the GWAS showing one SNP with genome-wide significant evidence of association in individuals of African genetic ancestry (bold) and top significantly associated SNP from prior GWAS of individuals of European genetic ancestry and replicated in this study. Frequency of the effect alleles in 1000 Genome populations are including for comparison. OR: Odds Ratio; Chr: Chromosome; GRCh38: Genome Reference Consortium Human Build 38. C: Cytosine; T: Thymine; * Indicates markers that were significantly associated in individuals of European ancestry (Klein et al., 2018).

| RsNumber | Chr:Position<br>(GRCh38) | Effect Allele/<br>Non-Effect<br>Allele | OR Effect<br>Allele | P Value | Effect Allele Frequency | | $r^2$ with<br>rs383009 | Effect Allele Frequency in<br>1000 Genome populations | |
| --- | --- | --- | --- | --- | --- | --- | --- | --- | --- |
|  |  |  |  |  | Cases | Controls |  | AFR | EUR |
| **rs383009** | **5:1327736** | **T/C** | **1.45** | **1.24 x 10<sup>-08</sup>** | **0.557** | **0.464** | **1.000** | **0.532** | **0.411** |
| **rs401681\*** | 5:1321972 | T/C | 1.32 | 2.28 x 10<sup>-05</sup> | 0.644 | 0.572 | 0.664 | 0.604 | 0.441 |
| **rs2736098\*** | 5:1293971 | T/C | 0.72 | 1.80 x 10<sup>-03</sup> | 0.095 | 0.129 | 0.107 | 0.066 | 0.235 |
| **rs35226131\*** | 5:1295258 | T/C | 1.50 | 1.07 x 10<sup>-01</sup> | 0.020 | 0.014 | 0.003 | 0.0098 | 0.036 |

### Polygenic Risk Score

In addition to examining known GWAS hits individually, we examined the fit of a weighted polygenic risk score (PRS) derived from the European ancestry population. A PRS was constructed for each individual by summing the number of risk alleles carried for all established pancreatic cancer risk loci identified by GWAS (two-sided P value < 5 x 10^-8^) previously published by Klein *et al*. [6], weighted by their estimated effect size. Individuals included in this study were grouped by PRS percentiles, and the association of this grouped PRS with pancreatic cancer was estimated using logistic regression with the middle quintile (40-60%) as the reference.

## Results

### Study Population

An overview of the study population is presented in **Table 1** and by study cohort, age and sex in **Supplemental Table 1.** Sex was not associated with case-control status (P value = 0.4). While we aimed to select controls of similar age to cases, there was some enrichment of cases in the extreme age groups (<55 and >75 years old, P value < 0.01, **Table 1**).

### Admixture Mapping

We evaluated the genetic contributions of local inferred African ancestry in relation to PDAC using admixture mapping. Three genome-wide significant regions were identified with higher inferred African ancestry proportions in PDAC cases (**Figure 1, bottom, Supplemental Table 3**) compared with controls. The first was observed at chr5q33.3 (chr5:159,677,942-159,685,714) with a higher proportion of African ancestry in cases (0.75) compared with controls (0.70) with an OR of 1.37 (95%CI = 1.19-1.57, P value= 4.41 x 10^-6^), **Figure 2**. Genes in this region include EBF transcription factor 1 (*EBF1*), RING Finger protein 145 (*RNF145*), and *Y_RNA* (**Figure 2**) among others.

**Figure 1.**
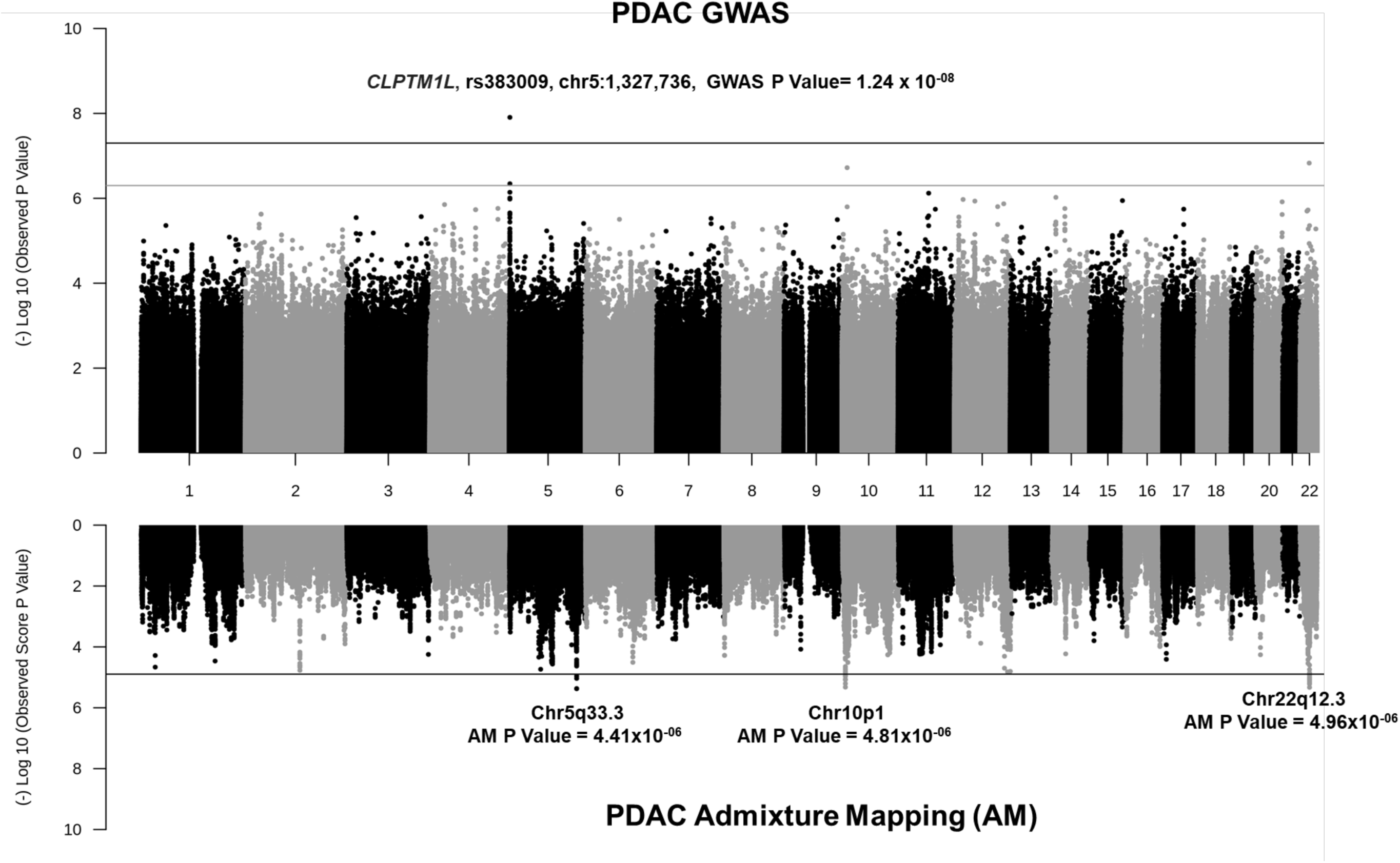
Miami plots summarizing the results of the genome-wide association analysis (“PDAC GWAS”, upper panel) and admixture mapping results (“PDAC Admixture mapping (AM)”, lower panel) in individuals of African genetic ancestry. Y and X axes correspond to (-) Log10 transformed P values for each analysis and the positions in chromosomes 1 to 22 based on GRCh38, respectively. The upper panel shows the results of the SNP based association and each point corresponds to the P value for a genetic autosomal variant. The black line represents the level of genome-wide significance (P value = 5 x10^-8^), and the gray line represents a suggestive association (P value = 5 x 10^-7^). Variants in 5p15.33 exceed the significance threshold (**Table 2**). Lower panel represents the results of the admixture mapping comparing the proportion of African vs European ancestry in each genomic regions in PDAC cases and controls. Each point corresponds to the P value for a genomic segment. The horizontal black line indicates the significance threshold (P value=1.27×10^-5^). Regions in chr5q33.3, chr10p1 and chr22q12.3 exceed the significance threshold **(Supplementary Table 3**).

**Figure 2.**
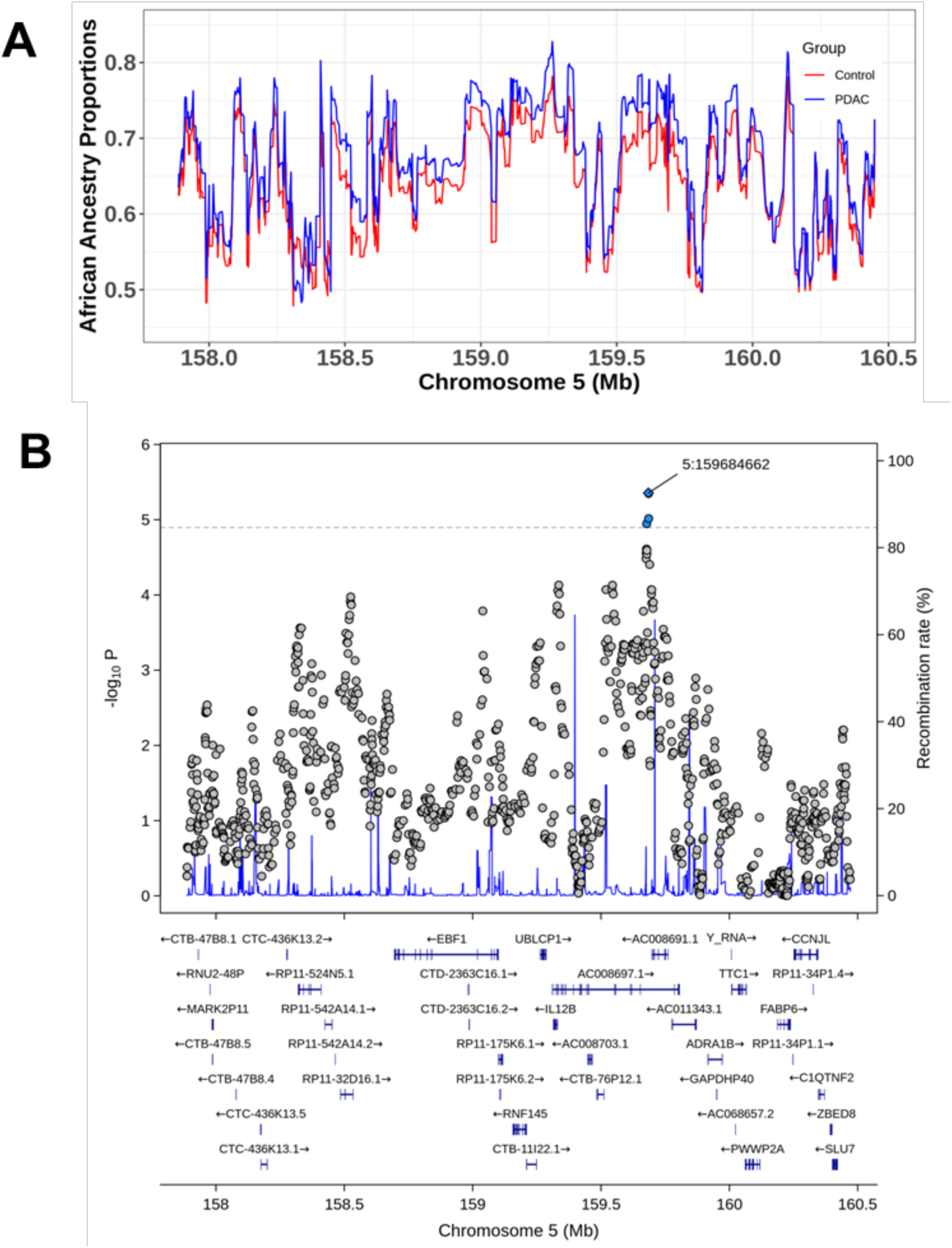
Results of the admixture mapping in chr5q33.3 region in the African ancestry population. Panel A shows the proportion of African ancestry in PDAC cases and controls by chromosome position. Lines correspond to local ancestry proportions in cases (blue) and controls (red) graphed as a function of the genomic coordinates in the region (GRCh38 assembly). Panel B presents the results of the admixture mapping. In this panel, each point represents an admixture genomic segment plotted with its P value as a function of genomic position. Segments with genome-wide significance are colored light blue. Recombination rates are estimated from UCSC database using the recomb1000GAvg track..The horizontal dashed line represents the significance level of the admixture mapping analysis (P value =1.27×10^-05^).

The second locus was a 52kb region located on chr22q12.3 (chr22:32,943,180-32,996,033). The top associated segment had an OR of 1.36 for PDAC when comparing African ancestry to European ancestry (95%CI = 1.18-1.56, P value= 4.96 x 10^-6^), **Supplemental Table 3, Figure 3**. African ancestry proportion in this region in cases varied from 0.65 to 0.70 and was significantly higher than in controls (0.57-0.61), **Figure 3, Panel A**. This region contains several genes including Synapsin III, transcript variant IIIc (*SYN3*), BPI fold containing family C (*BPIFC*), Large 1, TIMP metallopeptidase inhibitor 3 (*TIMP3*) and F-box protein 7 (*FBX07*) as presented in **Figure 3, Panel B**.

**Figure 3.**
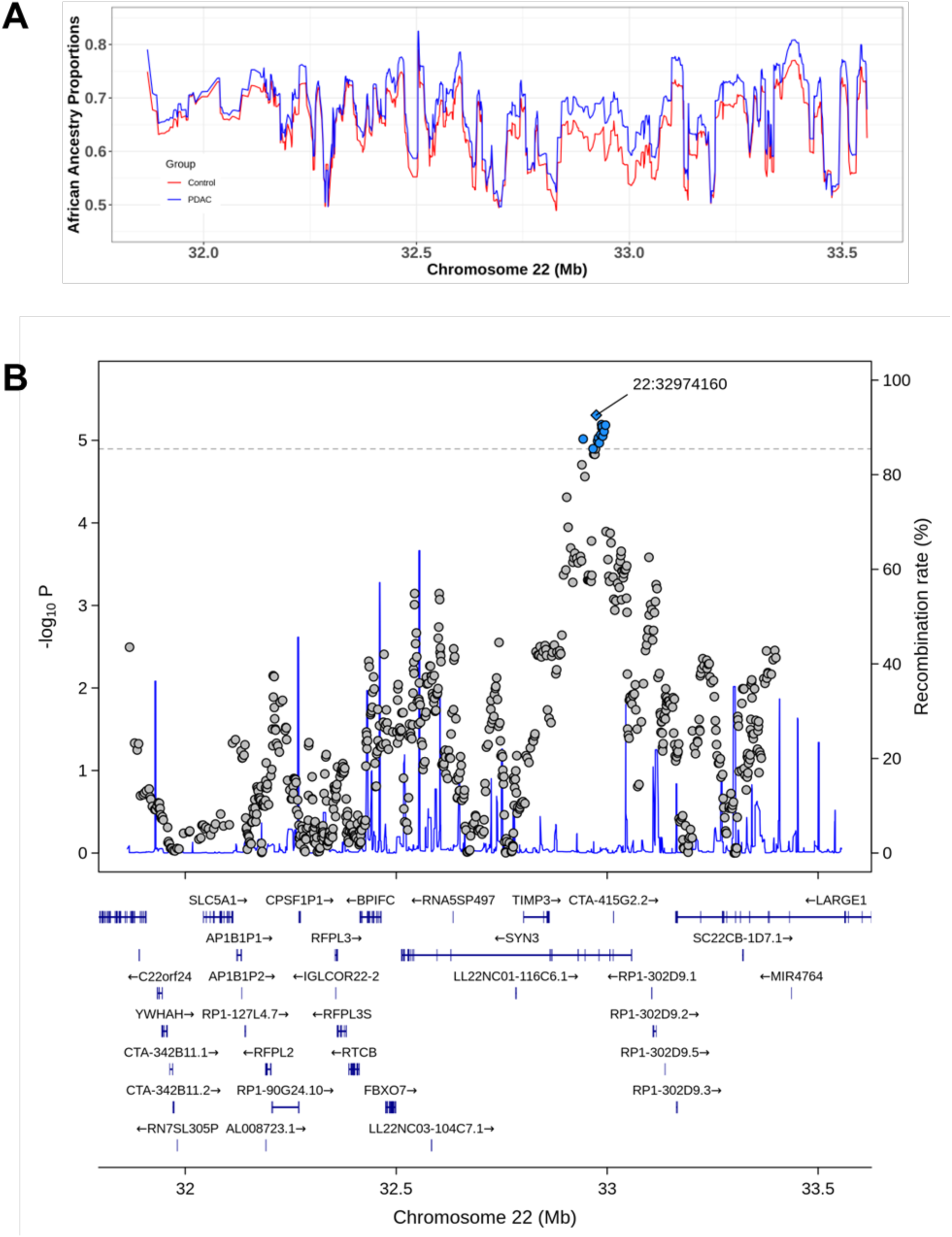
Results of the admixture mapping in the chr22q12.3 region in the African ancestry population. Panel A shows the proportion of African ancestry in PDAC cases and controls by chromosome position. Lines correspond to local ancestry proportions in cases (blue) and controls (red) graphed as a function of the genomic coordinates in the region (GRCh38 assembly). Panel B presents the results of the admixture mapping. In this panel, each point represents an admixture genomic segment plotted with its P value as a function of genomic position. Segments with genome-wide significance are colored light blue. Recombination rates are estimated from UCSC database using the recomb1000GAvg track.The horizontal dashed line represents the significance level of the admixture mapping analysis (P value =1.27×10^-05^)

The third significant locus was at chr10p1 (7.8kb, chr10:6,620,063-6,627,884; OR = 1.34, 95%CI = 1.19-1.51, P value = 4.8×10^-6^). This region contains *RP11-554I8.1* and long intergenic non-protein coding RNA 2648 (*LINC02648),* **Figure 4, Panel B.** Interestingly, in this region, the proportion of local African ancestry in the total sample was lower than the overall global ancestry (∼0.6), but still differed significantly between cases and controls (∼0.63 vs. 0.56), **Figure 4, Panel A**.

**Figure 4.**
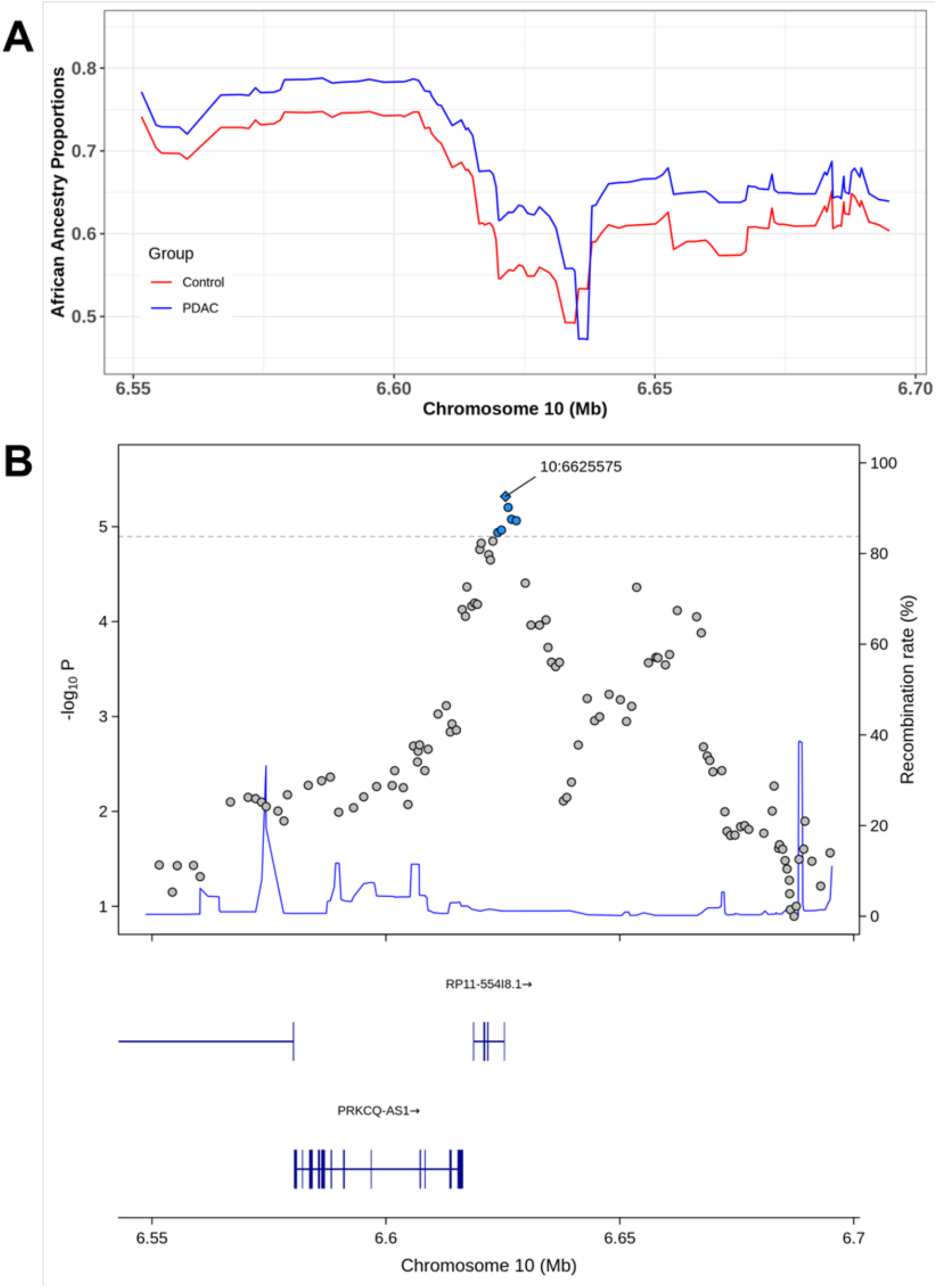
Results of the admixture mapping at chr10p14 region in the African ancestry population. Panel A shows the proportion of African ancestry in PDAC cases and controls by chromosome position. Lines correspond to local ancestry proportions in cases (blue) and controls (red) graphed as a function of the genomic coordinates in the region (GRCh38 assembly). Panel B presents the results of the admixture mapping. In this panel, each point represents an admixture genomic segment plotted with its P value as a function of genomic position. Segments with genome-wide significance are colored light blue. Recombination rates are estimated from UCSC database using the recomb1000GAvg track. The horizontal dashed line represents the significance level of the admixture mapping analysis (P value =1.27×10^-05^)

### Genome Wide SNP Association Analysis

We observed genome-wide significant evidence of association on chromosome 5p15.33 (**Figure 5)**. The SNP with the lowest P value was rs383009 (C>T), with per allele OR of 1.45 for PDAC (95%CI = 1.27-1.64, P value = 1.24 x 10^-8^) for the T allele compared to the C allele **(Figure 5**, **Table 2).** This genomic region is strongly associated with PDAC in prior GWAS of European ancestry [6]. The top SNPs in prior GWAS were rs2736098, rs35226131, and rs401681 [6] (**Supplemental Table 2),** and these were replicated in this study (**Figure 5 panel B and Table 2**). In conditional analysis, the lead SNP in this region (rs383009) fully accounted for the signal as no other SNPs provided evidence of association after adjusting for this SNP (P value > 0.05). To evaluate the potential effect of admixture on this association, we conditioned the SNP-based analysis on local ancestry. We found that the overall statistical significance decreased slightly, but the OR remained similar (rs383009 OR = 1.41, 95%CI = 1.22-1.63, P value = 2.01 x 10^-06^).

**Figure 5.**
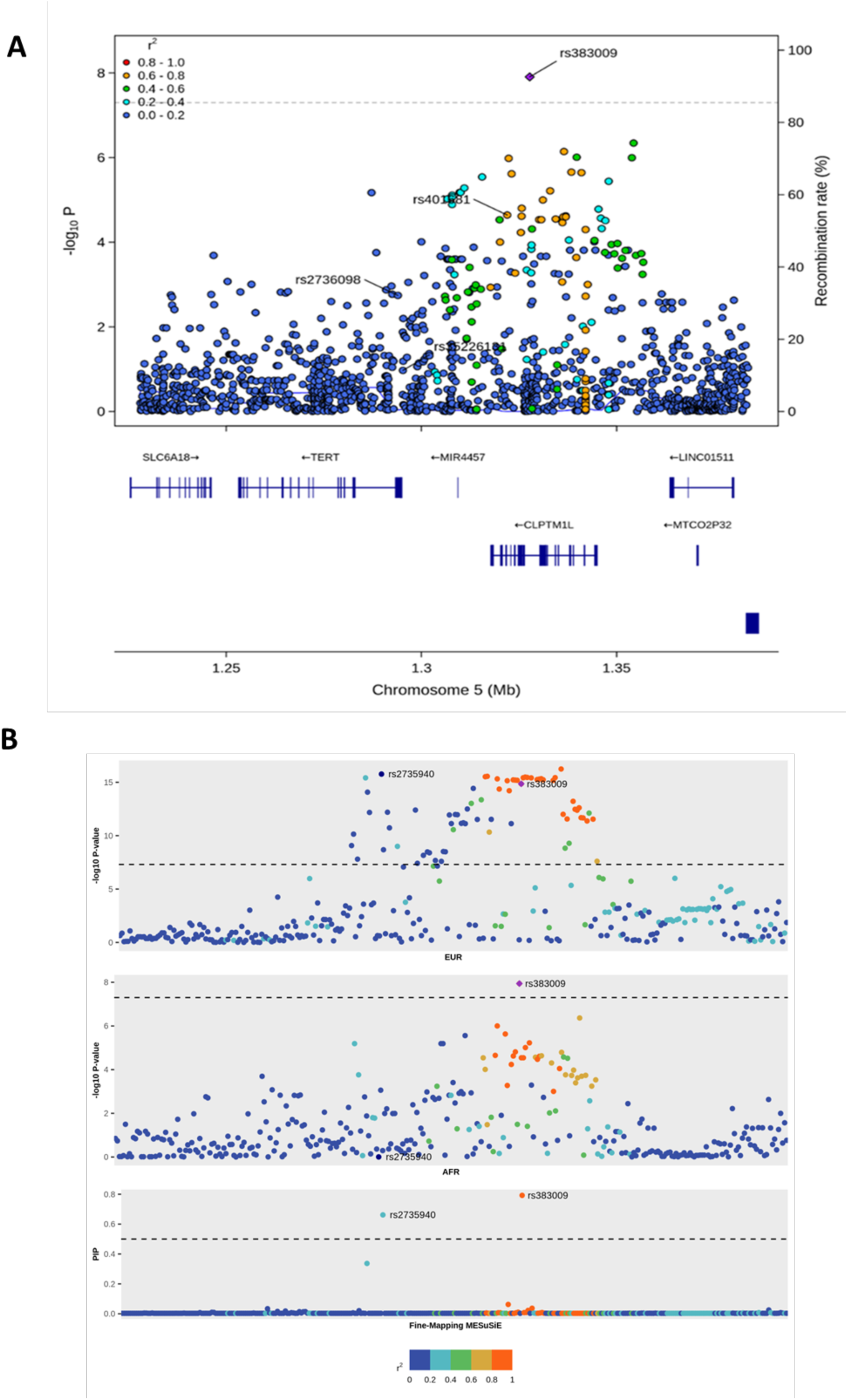
Locus zoom plots presenting the results of the SNP based association analysis at chr 5p15.33 in individuals of African genetic ancestry (Panel A) and multiancestry MESuSiE fine-mapping (Panel B). In Panel B, the three graphs from up to down represent results of the GWAS in European (EUR), African (AFR) ancestry groups and trans ancestral fine-mapping (MeSuSiE). Variants with PIP > 0.5 are annotated in the three graphs of Panel B. The X-axes for both panels correspond to the genomic position based on the GRCh38 assembly and the Y axes corresponds to the (-) Log P value of the GWAS as well and the PIP of the fine-mapping as annotated in each graph. Horizontal lines represent the significance threshold for each analysis (P value = 5 x 10^-08^ for GWAS and PIP>0.5 for MESuSie fine-mapping). Recombination rates in Panel A are estimated by using the recomb1000GAvg track from the UCSC database and genes definitions (represented as blue boxes) are based on USCS GENCODE Genes track (version 46, May 2024). In both panels, every dot represents an SNP colored based on *r^2^* values between each SNP in the region and the lead variant with PIP > 0.5 (rs383009, purple diamond) as described in the respective legends.

Of the genomic regions associated with PDAC risk in prior GWAS studies, we replicated three regions on 7q32.3, 8q24.21, and 17q25.1. In each of these regions, the same previously identified allele was associated with an increased risk of PDAC and the ORs were consistent with previous results (**Supplemental Table 2)**. In addition to those loci, prior studies have demonstrated a strong association with both SNPs in the *ABO* region, as well as with genotype derived ABO blood type [27]. We did not observe a statistically significant association with rs505922, the lead SNP in the *ABO* region on 9q34.2 (OR = 1.13, 95%CI = 0.98 - 1.29, P value = 0.08).

To explore this further, we constructed genotyped derived blood groups and tested for association with PDAC (**Supplemental Table 4**). In this analysis, we observed non-O blood group associated with an increased risk of PDAC (OR = 1.27, 95%CI = 1.05-1.53, P value = 0.01), consistent with prior studies.

### Fine-Mapping

Given the large number of associated variants in these GWAS regions, we conducted fine-mapping leveraging differences in LD patterns across ancestries and estimated the posterior inclusion probability (PIP) that a given variant “causal”, with higher probability indicating stronger statistical evidence supporting causality. In the GWAS-associated 5p15.33 locus, our multi-ancestral fine-mapping analysis covered a 159 kb region from chr5:1,224,719-1,383,931 and detected two variants (rs2735940 and rs383009) as potentially causal (PIP> 0.5). These two variants are located 31kb apart in the genes encoding telomerase reverse transcriptase (*TERT)* and lipid scramblase, also named cleft lip and palate transmembrane protein 1-like protein (*CLPTM1L)* as shown in **Figure 5**. Of note, rs383009 (chr5:1,327,736:C:T) is the SNP with the smallest P value in this current case-control analysis (**Supplemental Table 2 and Figure 5, Panel B**). This SNP is an intronic variant in *CLPTM1L* provided evidence of a shared effect in both European and African ancestry individuals (PIP=0.79), as displayed in **Figure 5, Panel B**. Comparison of the effect sizes of the GWAS analyses indicated that the odds for PDAC in individuals carrying the effect allele T vs allele C, was statistically significantly higher (P value=0.014) in individuals of African ancestry (OR=1.45, 95%CI=1.27-1.64) than that observed previously in European ancestry populations(OR=1.18, 95%CI=1.13-1.23). Interestingly, the allele frequencies are similar in these cohorts (T allele frequency European =0.46 and in our African GWAS=0.51). A second potentially causal variant, rs2735940 (chr5:1,296,371:A:G) was detected. It was associated with PDAC in European ancestry population (G allele OR= 1.19, 95%CI=1.14-1.24, PIP=0.66). However, no effect of this SNP was observed in African ancestry populations (OR=1.00, 95%CI=0.87-1.13, P value=0.99, G allele frequency in European=0.49 and African = 0.48), **Figure 5, Panel B**. Similar to rs383009, the effect of rs2735940 was also significant different between these populations (P value = 0.011)

When analyzing additional regions of genome-wide significant evidence of association in prior studies (**Supplementary Table 2**), we detected variants with a significant posterior inclusion probability at 1q32.1,1p36.33, 7p14.1, 7q32.3, 7p12, 8q24, and 13q22.1 **(Supplementary Figures 4-10).** In the chr1q32.1 region, there was evidence to support rs2821365 (chr1:200,041,800:A:G) as a causal variant (PIP=0.82) with effect sizes of OR=1.21 (95%CI=1.17-1.29, P value=1.67×10^-16^) in European ancestry individuals and OR=1.17 (95%CI=1.01-1.34, P value = 0.029) in individuals of African ancestry (**Supplemental Figure 4**). This variant is located in intron 2 of the nuclear receptor subfamily 5 group A member 2 *(NR5A2*) gene, which is abundantly expressed in pancreas. *NR5A2* haploinsufficiency has been associated with chronic pancreatitis, pancreatic, and gastrointestinal cancer [28]. Similarly, at 1p36.33, the putative causal variant rs13303160 (chr1:966,179:G:A, PIP=0.76) with shared effect in both populations (OR European ancestry= 0.83, 95%CI=0.76-0.90, P value = 2.59 x 10^-5^, OR African ancestry= 0.78, 95%CI=0.68-0.90, P value = 8 x 10^-4^) is located near *NOC2L* which encodes nucleolar complex protein 2 homolog (**Supplemental Figure 5**) [25] and *KLHL17* which encodes Kelch-like protein 17.

In region 7p12, rs73328512 (chr7:47,448,199:A:G) was the putatively causal SNP in our analysis as shown in **Supplemental Figure 6** (PIP = 0.77). This variant has a shared and consistently observed effect size in the African (G allele OR=0.87, 95%CI = 0.74-1.01, P value = 0.076) and European ancestry populations (G allele OR = 0.82, 95%CI = 0.77-0.88, P value = 9.7×10^−9^, **Supplemental Table 2**) and is located upstream of the tensin-2 (*TSN2)* gene.

In the 7p14.1 region, where the lead SNP in the European ancestry GWAS was rs17688601, we identified rs12701838 (chr7:40,837,874:A:G) as the putative causal variant in the region (PIP=0.51), **Supplementary Figure 7.** A decreased risk of PDAC was associated with the G vs the A allele in European ancestry individuals (OR G allele = 0.86, 95%CI = 0.83-0.91, P value = 2.40 x 10^-9^) but no association was observed in African ancestry individuals (OR = 0.99, 95%CI = 0.81-1.21, P value = 0.96) which is consistent with its ancestry specific effect. This variant, located in an intron of the succinyl-CoA:glutarate-CoA transferase (*SUGCT)* gene, is also an eQTL for the genes inhibin beta A chain (*INHBA*) and INHBA antisense RNA 1 (*INHBA-AS1*). The G allele was associated with higher levels of INHBA in tibial nerve (1.6 x 10^-30^), testis (2.1×10^-6^) as well as pancreas (1.5×10^-6^) in GTEx.

At 7q32.3, rs28651880 (chr7:130,992,183:C:T) was the most likely causal SNP in both European and African ancestry populations in our fine-mapping analysis (PIP=0.54), see **Supplemental Figure 8**. In our African ancestry GWAS analysis, we observed a lower risk of PDAC for the C allele compared to the T allele (OR = 0.77, 95%CI = 0.65-0.91, P value = 0.0023) which was consistent with the previously reported effect in European populations (OR = 0.80, 95%CI = 0.76-0.85, P value = 2.06 x10^-13^). Furthermore, the C allele has been associated with higher expression of the long intergenic non-protein coding RNA, p53 induced transcript gene (*LINC-PINT*) compared to the T allele in different tissues of GTEx database [29].

**Supplemental Figure 9** shows the fine-mapping results at chr8q24.21. Our analysis indicated that rs10094872 (chr8:127,707,639:A:T) was a potentially causal SNP (PIP=0.99). This SNP is ∼ 28 kb upstream of the *MYC* proto-oncogene. Allele T of the variant confers an OR =1.21 (95%CI = 1.06-1.40, P value= 0.0052) and OR = 1.15 (95%CI = 1.10-1.20, P value = 5.33×10^-11^) in African and European ancestry populations, respectively. The effect of this variant is shared across both populations. Similarly, at 13q22.1, rs9573166 (chr13:73,351,949:G:A, PIP = 0.85), had a shared effect (**Supplemental Figure 10)**. In both populations, the effect allele A conferred a lower risk of PDAC compared with the G allele, OR = 0.79 (95%CI = 0.74-0.85, P value = 6.3 x10^-12^) in European and a non-statistically significant OR = 0.85 (0.7-1.03, P value = 0.11) in African ancestry populations. Notably, there are marked differences in the frequency of the effect allele between these populations European (0.42) and African(0.12) ancestry populations. Interestingly, this same signal was identified in a prior transcriptome-wide association study in European ancestry populations [30] and in a functional investigation at this locus [31] where two genes, DIS3 exosome endoribonuclease and 3’-5’ (*DIS3)* and KLF transcription factor 5 (*KLF5)* were identified as likely target genes [31].

For the remaining GWAS regions and admixture mapping associated regions, no putative causal variants were identified **(Supplemental Table 2).**

### Polygenic Risk Score

We evaluated the performance of a previously developed 22-SNP polygenic risk score, based on European ancestry GWAS findings [6] in our inferred African ancestry individuals. The PRS was associated with PDAC risk, but effect sizes were generally lower across strata (**Supplemental Table 5**) compared with those observed in European ancestry populations [6]. For example, the odds ratio comparing the highest PRS decile to the 40^th^-60^th^ percentile bin was 1.45 (95% CI=1.02-2.07) in our African ancestry sample, as compared to 2.20 (95% CI=1.83–2.65) estimated in a previous European ancestry analysis [6].

## Discussion

Prior genomic studies have been limited by the relatively small number of PDAC cases available among self-reported Black individuals (<9,000 annually in the US), which has hindered the discovery of genetic variants relevant to different ancestries. To address this gap, we combined the resources of two large consortia, the Pancreatic Cancer Cohort and Pancreatic Cancer Case Control consortia, assembling one of the largest case-control studies of individuals with African ancestry to date.

Our study included 1,030 PDAC participants and 889 matched controls from the US and Canada. Participants were predominantly admixed, representing a broad spectrum of African ancestry proportions. We estimated genome-wide inferred ancestry as well as local ancestry at specific genomic regions to compare cases and controls. Using this approach, we identified three genomic regions in which PDAC risk was associated with increased ancestry at specific loci, highlighting candidate regions for future functional and genomic studies.

Importantly, the association signals in these regions were observed at the level of local ancestry segments, rather than specific allelic variants; as such, allele-based fine-mapping did not identify specific putative causal single-nucleotide variants. However, each of these regions contains candidate genes that may be relevant to PDAC pathogenesis. For example, the chr5q33.3 region contains *EBF1*, a transcriptional repressor of the telomerase catalytic subunit (*TERT*), a likely target gene at the established PDAC risk locus on chr5p15.33. Inactivation of *EBF1* is a major cause of *TERT*’s upregulation in gastric cancer [32]. This region also contains *RNF145* a member of the RING finger protein family frequently dysregulated in human malignancies, including hepatocellular carcinoma [33]. Overexpression of *RNF145* is associated with more aggressive ovarian cancer [34] and progression of oral squamous cell carcinoma [35]. Another candidate gene in this region is *SNORA68*, encoding small nucleolar RNA, H/ACA box 68, which has been implicated in triple-negative breast cancer [36], and has been proposed as a proposed biomarker for esophageal cancer and non-small cell lung cancer [37].

Similarly, genes in the chr22q12.3 region identified through admixture mapping are relevant to cancer biology. *TIMP3* is a key regulator of extracellular matrix remodeling and has been implicated in the progression of various cancers, including PDAC [38]. Likewise, *RFPL3S* has been proposed as a prognostic biomarker in lung cancer [39] and a suppressor and prognostic predictor in testicular cancer [40].

The GWAS-associated region at chr5p15.33 is a well-established multi-cancer risk locus with at least 10 independent signals reported across different cancers, and several populations [6, 8, 41, 42]. Notably, the risk-increasing alleles vary across cancer types [43, 44]. In our study of African ancestry cases and controls, the PDAC-associated SNP with the smallest P value (rs383009) was not the same as the lead SNP in previously-reported European ancestry GWAS (rs401681), although there is strong linkage disequilibrium between these SNPs in the 1,000 Genomes Project AFR and EUR reference panels (*r^2^*=0.84 and *r^2^*=0.89, respectively) ), indicating that this locus is not ancestry specific. Moreover, rs383009 reached genome-wide significance in the European ancestry GWAS and our multi-ancestral fine-mapping suggests this variant has a shared effect on both populations. This intronic variant in *CLPTM1L* has been described as a *cis*-regulatory eQTL for both *TERT* and *CLPTM1L* [45] and both genes were identified in a transcriptome-wide association study for PDAC [30, 45].

Importantly, the effect size of the potential causal variants (rs383009 and rs2735940) were different between the European ancestry GWAS [6] population and the inferred African ancestry population. The observed effect size for rs383009, was higher in African Ancestry OR=1.45, 95%CI=1.27-1.64) compared to European ancestry populations (OR=1.18, 95%CI=1.13-1.23). Conversely, rs2735940 was only associated in the European ancestry population (OR for the G allele vs A allele = 1.19, 95%CI=1.14-1.24) but not in the African ancestry populations (OR=1.00, 95%CI=0.87-1.13). At 1p36.33, our fine-mapping analysis identified rs13303160 as the most likely causative variant, shared across different ancestries. This finding supports our earlier fine-mapping analysis and functional studies conducted in populations of European ancestry [46]. In our prior study, rs13303160 was identified as a functional SNP, with the risk-increasing allele associated with lower KLHL17 mRNA expression. Additionally, lower KLHL17 expression was associated with pro-inflammatory pathways, suggesting its important role in mediating risk at this locus [46]. Overall, our current findings highlight the relevance of this variant across diverse populations.

The fine-mapping analysis in the 7p14.1 region identified rs12701838 as a putative causal variant; however, the evidence was limited to individuals of European ancestry. The G allele was associated with a decreased risk of pancreatic cancer; this variant is also associated with higher expression of inhibin subunit beta A (*INHBA*, which encodes a subunit of Activin A and Inhibin A) and INHBA antisense RNA 1 (*INHBA-AS1*) in GTEx. Activins are members of the TGF-Beta superfamily, and Activin A has been associated with cancer cell proliferation, migration and invasion [47]. High Activin A levels are associated with pancreatic cancer cachexia by increasing visceral adipose tissue wasting [48], and high expression of Activin A in the tumor adjacent stromal cells has been associated with a poor prognosis [49]. Interestingly, the lower risk is in carriers of the rs12701838-G allele, which is associated with higher INHBA levels [26]. As such, additional work is needed to understand the observed association at this locus.

Collectively, these findings provide important context for interpreting population differences in PDAC susceptibility. Although we assembled one of the largest cohorts of individuals with African ancestry and PDAC to date, our sample size remains limited compared to GWAS studies in European populations, which may limit power to detect weaker or ancestry-specific effects.

Differences in allele frequencies alone do not fully account for the observed variation in pancreatic cancer risk across populations. Rather, the combined results from the admixture mapping, GWAS, and fine-mapping indicate that while some PDAC risk regions are shared among populations, the strength of the genetic associations at these loci may differ significantly. At the well-established chr5q15.33 locus, our analyses support a largely shared underlying signal between genetically inferred African ancestry and European ancestry populations reinforcing the concept that shared biologic pathways may underlie PDAC susceptibility across populations, although the architecture of these genetic associations may vary. This emphasizes the importance of performing genetic studies in diverse populations, not only to aid locus discovery and fine-mapping resolution, but also to improve the development and application of polygenic risk scores and to better characterize the heterogeneity in effect sizes and risk across ancestry groups. Importantly, while genomic studies are essential to understand inherited susceptibility, lifestyle and environmental factors also contribute significantly to PDAC risk. By integrating insights from genetics with knowledge of lifestyle influences, we can enhance our understanding of pancreatic cancer and advance prevention and treatment strategies for all populations.

## Data availability

Data from this study will be made available in dbGAP under phs003492, phs000206 and phs000648 under controlled access.

