## Supplemental Materials for "Novel Genetic Risk Loci for Pancreatic Ductal Adenocarcinoma Identified in a Genome-wide Study of African Ancestry Individuals"

### Population Supplemental Tables and Figures

**Supplemental Table 1**. Demographic characteristics of the study participants by affection status, site of recruitment, and study cohort.

| Study Name | Cases n (%) | Controls n (%) | Study Design | Source | | Reference |
|---|---|---|---|---|---|---|
| | | | | Cases | Controls | |
| **American Cancer Society - Cancer Prevention Study -II** | 1(20) | 4(80) | Cohort | Self-report & National Death Index with medical record and state cancer registries | | Calle et al PMID 11900235 |
| **Columbia University** | 24(86) | 4(14) | Case/Control | Clinic | | |
| **Dana Farber Cancer Institute** | 30(100) | 0 | Case/Control | Clinic | | |
| **Fox Chase Cancer Center** | 17(43) | 23(58) | Case/Control | Clinic/Biosample Repository | Healthy Volunteers | Sridharan et al. PMID: 39954184 |
| **Johns Hopkins Hospital** | 145(97) | 4(3) | Family Case Control | Clinic & Family Cohort | Spouse/Friend | |
| **Karmanos Cancer Center** | 0 | 198(100) | Cohort Controls | | | |
| **MD Anderson Cancer Center** | 213(93) | 15(7) | Case/Control | Clinic | Friends/Spouses of non-pancreatic cancer patients | Hassan et al PMC2423805) |
| **Multiethinic Cohort Study of Diet and Health** | 98(49) | 101(51) | Cohort | Hawaii Cancer Registry, Cancer Surveillance Program of Los Angeles County and California State Cancer Registry | | Kolonel et al PMID10695593 |
| **Memorial Sloan Kettering Cancer Center** | 8(62) | 5(38) | Case/Control | Clinic | | Olson SH et al PMID18031948) |
| **Mayo Clinic Molecular Epidemiology Case Control Study** | 57(86) | 9(14) | Case/Control | Clinic | Primary Care Patients | McWilliams et al PMC2652067) |
| **Moffit Cancer Center on behalf of the Florida Pancreas Collaborative** | 74(100) | 0 | Cohort | Clinic | | Permuth et al PMID33671939 |
| **New York University Women's Health Study** | 2(22) | 7(78) | Cohort | | | Zeleniuch-Jacquotte et al PMID 15540225) |
| **Prostate, Lung, Colorectal, Ovarian Cancer Screening Trial (PLCO)** | 16(9) | 156(91) | Cohort | Self-report with pathology report confirmation., linkage to cancer registries, and national death index. | | Black et al PMID PMID: 26435289 |
| **University of Toronto, Sinai Health** | 20(63) | 12(38) | Population Based Hospital and Cancer Registry | Clinic | Family Medicine Clinical Database | Eppel A et al PMID1752608 |
| **Southern Community Cohort Study** | 197(49) | 203(51) | Cohort | | | |
| **Women's Health Initiative** | 84(49) | 89(51) | Cohort | Self-report with medical records | | Rexrode KM et al PMC3714020) |
| **Yale** | 17(41) | 24(59) | Case/Control | Population Based Hospital and Cancer Registry | | Risch HA PMID 12837831) |
| **University of San Francisco** | 27(46) | 35(56) | Case/Control | Clinic | Clinic | Duell EJ et al PMID18559563 |

**Supplemental Table 2**. Genomic regions harboring genome-wide significant or marginal association signals with PDAC in European ancestry population and corresponding results obtained in the GWAS in individuals of African ancestry. In bold are loci that showed replication with a P value < 0.01. Acronyms: EAF: Effect Allele Frequency. GRCh38: Genome Reference Consortium Human Genome build 38.

| Chromosome region<br>Top SNP<br>Position (GRCh38)<br>Gene | Effect Allele / Non-Effect Allele | Statistics | European Ancestry GWAS (Klein et al 2018) | African Ancestry GWAS | PIP>0.5 in both GWAS<br>rsnumber<br>Position (GRCh38) |
|---|---|---|---|---|---|
| 1q32.1 | A/T | EAF cases;controls | 0.26;0.23 | 0.74;0.70 | rs2821365<br>1:200,041,800:A:G<br>PIP=0.82 |
| rs2816938 | | Imputation quality (info) | 0.998 | | |
| 200,016,240 | | OR (CI) | 1.21 (1.17 - 1.26) | 1.17 (1.01 - 1.35) | |
| NR5A2 | | P value GWAS | 3.362x10$^{-15}$ | 0.04 | |
| 1q32.1 | G/A | EAF cases;controls | 0.20;0.23 | 0.12;0.14 | |
| rs3790844 | | Imputation quality (info) | Genotyped | | |
| 200,038,304 | | OR (CI) | 0.81 (0.76 - 0.86) | 0.85 (0.70 - 1.03) | |
| NR5A2 | | P value GWAS | 7.619x10$^{-16}$ | 0.1 | |
| 1p36.33 | G/A | EAF cases;controls | 0.13;0.11 | 0.75;0.71 | rs13303160<br>1:966,179:G:A<br>PIP=0.75 |
| rs13303010 | | Imputation quality (info) | Genotyped | | |
| 959,193 | | OR (CI) | 1.20 (1.12 -1.29) | 1.15 (0.99-1.34) | |
| NOC2L | | P value GWAS | 0.00000073 | 0.06 | |
| 2p13.3 | C/T | EAF cases;controls | 0.30;0.28 | 0.18;0.16 | No variant detected |
| rs2035565 | | Imputation quality (info) | 0.999 | | |
| 67,392,524 | | OR (CI) | 1.12 (1.07 - 1.16) | 1.11 (0.94 - 1.31) | |
| ETAA1 | | P value GWAS | 0.000002555 | 0.23 | |
| 2p13.3 | G/T | EAF cases;controls | 0.30;0.28 | 0.18;0.17 | No variant detected |
| rs1486134 | | Imputation quality (info) | Genotyped | | |
| 67,412,637 | | OR (CI) | 1.12 (1.07 - 1.16) | 1.09 (0.93 - 1.29) | |
| ETAA1 (2236bp 3') | | P value GWAS | 9.797x10$^{-07}$ | 0.3 | |
| 3q29 | A/G | EAF cases;controls | 0.33; 0.37 | 0.27;0.25 | No variant detected |
| rs9854771 | | Imputation quality (info) | 1 | | |
| 189,790,682 | | OR (CI) | 0.89 (0.85 - 0.93) | 1.09 (0.93 - 1.27) | |
| TP63 | | P value GWAS | 1.14410x$^{-07}$ | 0.28 | |
| 5p15.33 | T/C | EAF cases;controls | 0.24;0.27 | 0.10;0.13 | |
| rs2736098 | | Imputation quality (info) | 0.921 | | |
| 1,293,971 | | OR (CI) | 0.83 (0.78 - 0.88) | 0.72 (0.59 - 0.88) | rs383009<br>5:1,327,736:C:T<br>PIP=0.79 |
| TERT | | P value GWAS | 5.803x10$^{-14}$ | **0.0018** | |
| 5p15.33 | T/C | EAF cases;controls | 0.02;0.03 | 0.02;0.01 | |
| rs35226131 | | Imputation quality (info) | 0.981 | | rs2735940 |

| | | | | | |
|---|---|---|---|---|---|
| 1,295,258 | | OR (CI) | 0.67 (0.53 - 0.81) | 1.50 (0.92 - 2.46) | 5:1,296,371:A:G PIP=0.661 |
| | | P value GWAS | 2.188x10$^{-08}$ | 0.11 | |
| 5p15.33 | T/C | EAF cases;controls | 0.49;0.44 | 0.64;0.57 | |
| rs401681 | | Imputation quality (info) | Genotyped | | |
| 1,321,972 | | OR (CI) | 1.19 (1.15 - 1.23) | 1.32 (1.16 - 1.51) | |
| CLPTM1L | | P value GWAS | 9.315x 10$^{-17}$ | **0.0000228** | |
| 7p14.1 | | EAF cases;controls | 0.25;0.27 | 0.07;0.07 | rs12701838 |
| rs17688601 | A/C | Imputation quality (info) | Genotyped | | 7:40,837,874:A:G PIP=0.51 |
| 40,827,064 | | OR (CI) | 0.88 (0.83 - 0.93) | 1.01 (0.78 - 1.31) | |
| SUGCT | | P value GWAS | 8.232x10$^{-08}$ | 0.93 | |
| 7q32.3 | | EAF cases;controls | 0.14;0.16 | 0.14;0.17 | rs28651880 |
| rs6971499 | C/T | Imputation quality (info) | Genotyped | | 7:130,992,183:C:T PIP=0.54 |
| 130995762 | | OR (CI) | 0.82 (0.76 - 0.88) | 0.79 (0.66 - 0.95) | |
| LINC-PINT | | P value GWAS | 4.323x10$^{-11}$ | **0.0108** | |
| 7p12.3 | | EAF cases;controls | 0.10;0.12 | 0.22;0.24 | rs73328512 |
| rs73328514 | T/A | Imputation quality (info) | 0.97 | | 7:47,448,199:A:G PIP=0.77 |
| 47,448,971 | | OR (CI) | 0.83 (0.77-0.88) | 0.88 (0.75 - 1.02) | |
| TNS3 | | P value GWAS | 4.35x10$^{-08}$ | 0.09 | |
| 8q24.21 | | EAF cases;controls | 0.39;0.37 | 0.33;0.29 | |
| rs10094872 | | Imputation quality (info) | 0.967 | | rs10094872 |
| 127,707,639 | T/A | OR (CI) | 1.14 (1.10 - 1.19) | 1.22 (1.06 - 1.40) | 8:127,707,639:A:T PIP=0.99 |
| MYC | | P value GWAS | 1.19x10$^{-09}$ | **0.0052** | |
| 8q24.21 | | EAF cases;controls | 0.24;0.26 | 0.55;0.56 | |
| rs1561927 | C/T | Imputation quality (info) | Genotyped | | |
| 128555832 | | OR (CI) | 0.89 (0.84 - 0.93) | 0.94 (0.82 - 1.07) | |
| MIR1208 | | P value GWAS | 6.178x10$^{-07}$ | 0.33 | |
| 8q21.11 | | EAF cases;controls | 0.41;0.43 | 0.14;0.15 | |
| rs2941471 | G/A | Imputation quality (info) | 1 | | No variant |
| 75,558,169 | | OR (CI) | 0.89 (0.86-0.94) | 0.88 (0.73,1.07) | |
| HNF4G | | P value GWAS | 0.000000473 | 0.21 | |
| 9q34 | | EAF cases;controls | | 0.37;0.35 | |
| rs505922 | C/T | Imputation quality (info) | Genotyped | | No variant |
| 133273813 | | OR (CI) | 1.27 (1.22 - 1.31) | 1.13 (0.98 - 1.29) | |
| ABO | | P value GWAS | 7.351x10$^{-27}$ | 0.08 | |
| 13q12.2 | | EAF cases;controls | 0.43;0.39 | 0.14;0.14 | |
| rs9581943 | A/G | Imputation quality (info) | Genotyped | | No variant |
| 27,919,860 | | OR (CI) | 1.16 (1.12 - 1.21) | 1.06 (0.88 - 1.28) | |
| PDX1 | | P value GWAS | 1.205x10$^{-12}$ | 0.55 | |
| 13q22.1 | C/T | EAF cases;controls | 0.43;0.37 | 0.87;0.85 | rs9573166 |

| | | | | | |
|---|---|---|---|---|---|
| rs9543325 | | Imputation quality (info) | Genotyped | | 13:73,351,949:G:A PIP=0.85 |
| 73342491 | | OR (CI) | 1.24 (1.19 - 1.28) | 1.12 (0.93 - 1.37) | |
| KLF5 and KLF12 | | P value GWAS | 1.216x10$^{-22}$ | 0.23 | |
| 16q23.1 | A/G | EAF cases;controls | 0.06;0.05 | 0.24;0.24 | No variant |
| rs7190458 | | Imputation quality (info) | Genotyped | | |
| 75,229,763 | | OR (CI) | 1.36 (1.26 -1.46) | 1.01 (0.87 - 1.17) | |
| BCAR1 | | P value GWAS | 7.092x10$^{-10}$ | 0.9 | |
| 17q25.1 | T/C | EAF cases;controls | 0.14;0.11 | 0.25;0.21 | No variant |
| rs11655237 | | Imputation quality (info) | 0.955 | | |
| 72,404,025 | | OR (CI) | 1.25 (1.19 - 1.31) | 1.28 (1.10 - 1.50) | |
| LINC00673 | | P value GWAS | 4.651x10$^{-12}$ | **0.00162** | |
| 17q25.1 | T/C | EAF cases;controls | 0.14;0.11 | 0.25;0.20 | |
| rs7214041 | | Imputation quality (info) | Genotyped | | |
| 72405335 | | OR (CI) | 1.25 (1.18 - 1.31) | 1.29 (1.11 - 1.51) | |
| LINC00673 | | P value GWAS | 6.581x10$^{-12}$ | **0.00127** | |
| 17q12 | A/G | EAF cases;controls | 0.21;0.23 | 0.06;0.07 | No variant |
| rs4795218 | | Imputation quality (info) | 0.95 | | |
| 37,718,512 | | OR (CI) | 0.88 (0.82 -0.93) | 0.89 (0.68,1.16) | |
| HNF1B | | P value GWAS | 0.000000273 | 0.39 | |
| 18q21.32 | T/C | EAF cases;controls | 0.17;0.18 | 0.26;0.29 | No variant |
| rs1517037 | | Imputation quality (info) | Genotyped | | |
| 59,211,042 | | OR (CI) | 0.87(0.82-0.93) | 0.84 (0.73,0.97) | |
| GRP | | P value GWAS | 0.000000881 | 0.02 | |
| 22q12.1 | T/C | EAF cases;controls | 0.17;0.16 | 0.05;0.04 | No variant |
| rs16986825 | | Imputation quality (info) | Genotyped | | |
| 28,904,318 | | OR (CI) | 1.16 (1.10 - 1.21) | 1.13 (0.84,1.53) | |
| ZNRF3 | | P value GWAS | 2.926x10$^{-07}$ | 0.41 | |

**Supplemental Table 3**. Regions with genome-wide significant evidence of association between local African ancestry and PDAC detected via admixture mapping. OR: Odds ratio, SE: Standard error, Chr: chromosome, GRCh38: Genome Research Consortium human build 38.

| Chr | Position (GRCh38) | Local African Ancestry proportion | | | OR | SE | P Value |
|---|---|---|---|---|---|---|---|
| | | Total sample (N=1,919) | Cases (n=1,030) | Controls (n=889) | | | |
| **5** | 159677942 | 0.70 | 0.73 | 0.67 | 1.35 | 0.07 | $1.13 \times 10^{-05}$ |
| **5** | 159684662 | 0.70 | 0.73 | 0.66 | 1.37 | 0.07 | $4.41 \times 10^{-06}$ |
| **5** | 159685551 | 0.70 | 0.73 | 0.66 | 1.37 | 0.07 | $4.47 \times 10^{-06}$ |
| **5** | 159685714 | 0.70 | 0.73 | 0.66 | 1.36 | 0.07 | $9.65 \times 10^{-06}$ |
| **10** | 6623909 | 0.60 | 0.63 | 0.56 | 1.32 | 0.06 | $1.16 \times 10^{-05}$ |
| **10** | 6624697 | 0.60 | 0.63 | 0.56 | 1.32 | 0.06 | $1.09 \times 10^{-05}$ |
| **10** | 6625575 | 0.59 | 0.62 | 0.55 | 1.34 | 0.06 | $4.81 \times 10^{-06}$ |
| **10** | 6626134 | 0.59 | 0.62 | 0.55 | 1.33 | 0.06 | $6.28 \times 10^{-06}$ |
| **10** | 6626868 | 0.59 | 0.62 | 0.55 | 1.33 | 0.06 | $8.37 \times 10^{-06}$ |
| **10** | 6627884 | 0.60 | 0.63 | 0.56 | 1.33 | 0.06 | $8.66 \times 10^{-06}$ |
| **22** | 32943180 | 0.62 | 0.65 | 0.58 | 1.34 | 0.07 | $9.64 \times 10^{-06}$ |
| **22** | 32966537 | 0.67 | 0.70 | 0.63 | 1.34 | 0.07 | $1.27 \times 10^{-05}$ |
| **22** | 32974160 | 0.64 | 0.67 | 0.60 | 1.36 | 0.07 | $4.96 \times 10^{-06}$ |
| **22** | 32977231 | 0.64 | 0.67 | 0.61 | 1.34 | 0.07 | $1.02 \times 10^{-05}$ |
| **22** | 32979153 | 0.65 | 0.68 | 0.61 | 1.35 | 0.07 | $9.34 \times 10^{-06}$ |
| **22** | 32981517 | 0.64 | 0.68 | 0.61 | 1.34 | 0.07 | $1.08 \times 10^{-05}$ |
| **22** | 32984776 | 0.61 | 0.65 | 0.58 | 1.34 | 0.07 | $8.54 \times 10^{-06}$ |
| **22** | 32986770 | 0.62 | 0.65 | 0.58 | 1.35 | 0.07 | $6.46 \times 10^{-06}$ |
| **22** | 32987434 | 0.61 | 0.65 | 0.58 | 1.35 | 0.07 | $7.06 \times 10^{-06}$ |
| **22** | 32988901 | 0.61 | 0.65 | 0.58 | 1.35 | 0.07 | $6.81 \times 10^{-06}$ |
| **22** | 32989705 | 0.61 | 0.65 | 0.57 | 1.34 | 0.07 | $8.90 \times 10^{-06}$ |
| **22** | 32992792 | 0.61 | 0.65 | 0.58 | 1.35 | 0.07 | $7.81 \times 10^{-06}$ |
| **22** | 32996033 | 0.61 | 0.65 | 0.57 | 1.35 | 0.07 | $6.55 \times 10^{-06}$ |

**Supplemental Table 4.** Association of ABO blood groups and PDAC risk in individuals of African Ancestry. OR: Odds ratio; CI: confidence interval; Ref: Reference category.

| ABO group | Cases n (%) | Controls n (%) | OR (95% CI) |
|---|---|---|---|
| **O** | 350 (33.9) | 351 (39.4) | Ref. |
| **Non-O** | 680 (66.0) | 538 (60.5) | 1.27 (1.05-1.53) |
| **Total** | 1030 | 889 | |

**Supplemental Table 5.** Performance of European Ancestry polygenic risk score in African Americans. Acronyms: PRS: polygenic risk score; OR: Odds ratio; CI: confidence interval; Ref: Reference category.

| PRS percentile | OR (95%CI) | P value | Cases, n (%) |
|---|---|---|---|
| **<10%** | 0.58 (0.41-0.82) | 0.002 | 80 (7.7%) |
| **10-<20%** | 0.60 (0.42-0.85) | 0.004 | 82 (7.9%) |
| **20-<40%** | 0.78 (0.59-1.04) | 0.090 | 189 (18.2%) |
| **40-<60%** | Ref. | Ref. | 213 (20.1%) |
| **60-<80%** | 1.13 (0.85-1.50) | 0.400 | 224 (21.7%) |
| **80-<90%** | 1.41 (0.96-2.01) | 0.060 | 122 (11.8%) |
| **>90%** | 1.45 (1.02-2.07) | 0.041 | 124 (12.0%) |

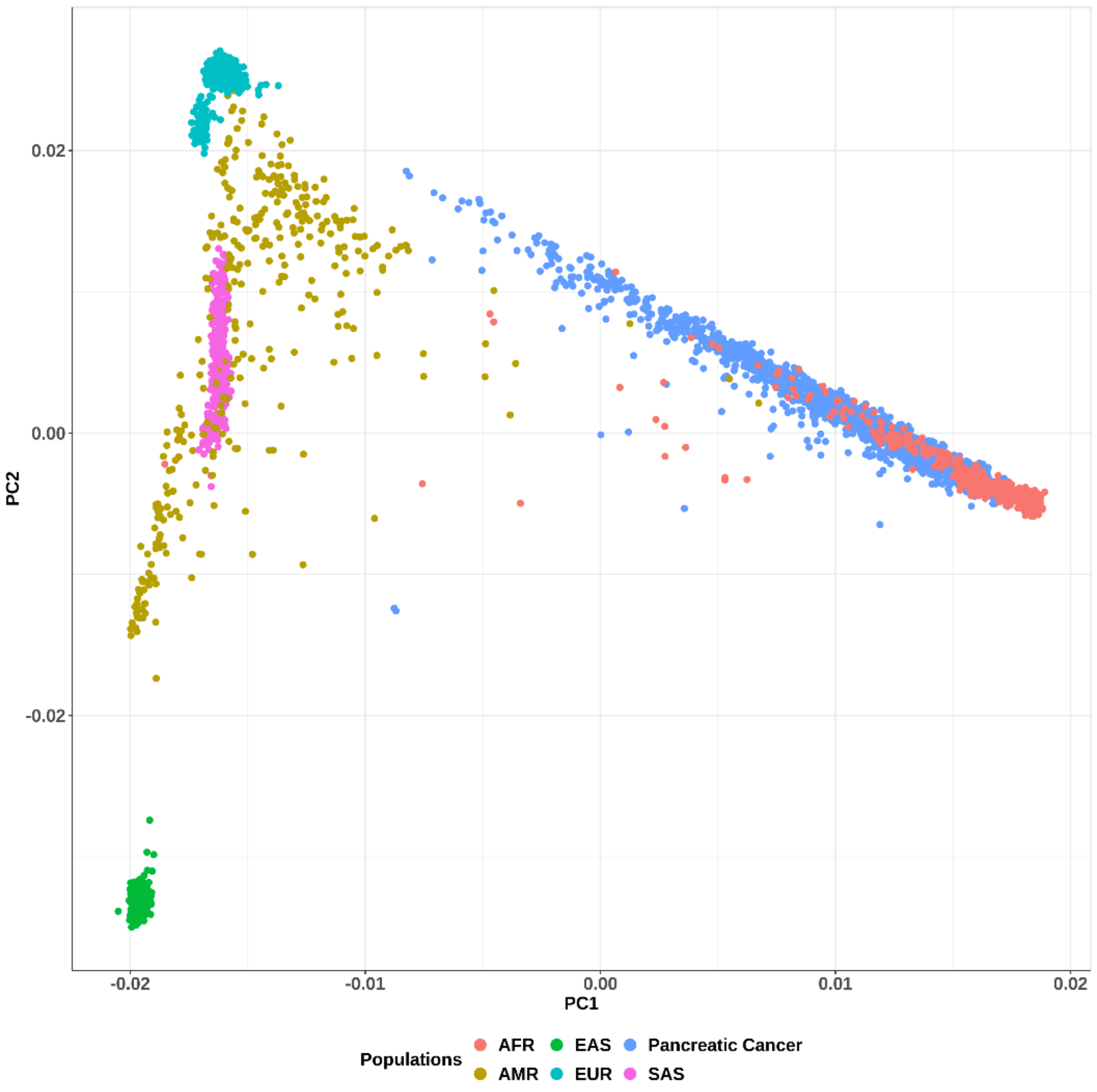


**Supplemental Figure 1.** Principal component analysis of the individuals diagnosed with PDAC and controls included in this study along with populations of 1000 Genomes project. Populations are color coded based on their genetically determined population ancestry groups as described in the legend. X and Y axes represent the first two principal components (PC1 and PC2) as determined from the ancestry-specific principal component analysis. Individuals of 1000 Genomes are categorized as Africans (AFR), East Asians (EAS), Europeans (EUR), Americans (AMR) and South Asians (SAS). PDAC cases and controls in the current study are graphed together and labeled as "Pancreatic cancer" group.

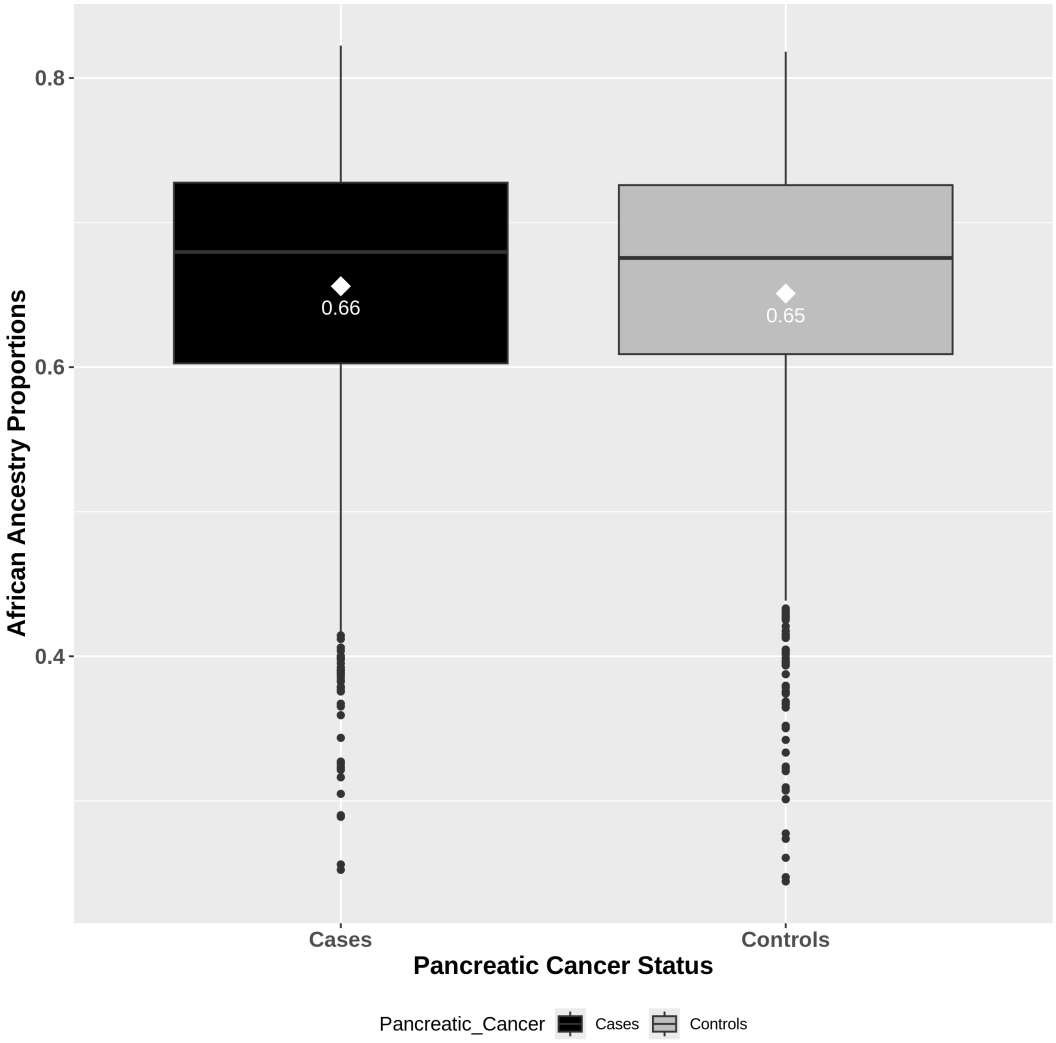


**Supplemental Figure 2.** Box plots of proportions of global genetically determined African ancestry proportions in individuals with PDAC and controls. The horizontal line represents the median of global African ancestry in each group, and the whiskers represent the 25-75 percentiles. The values inside the plot indicate the average proportions for each category.

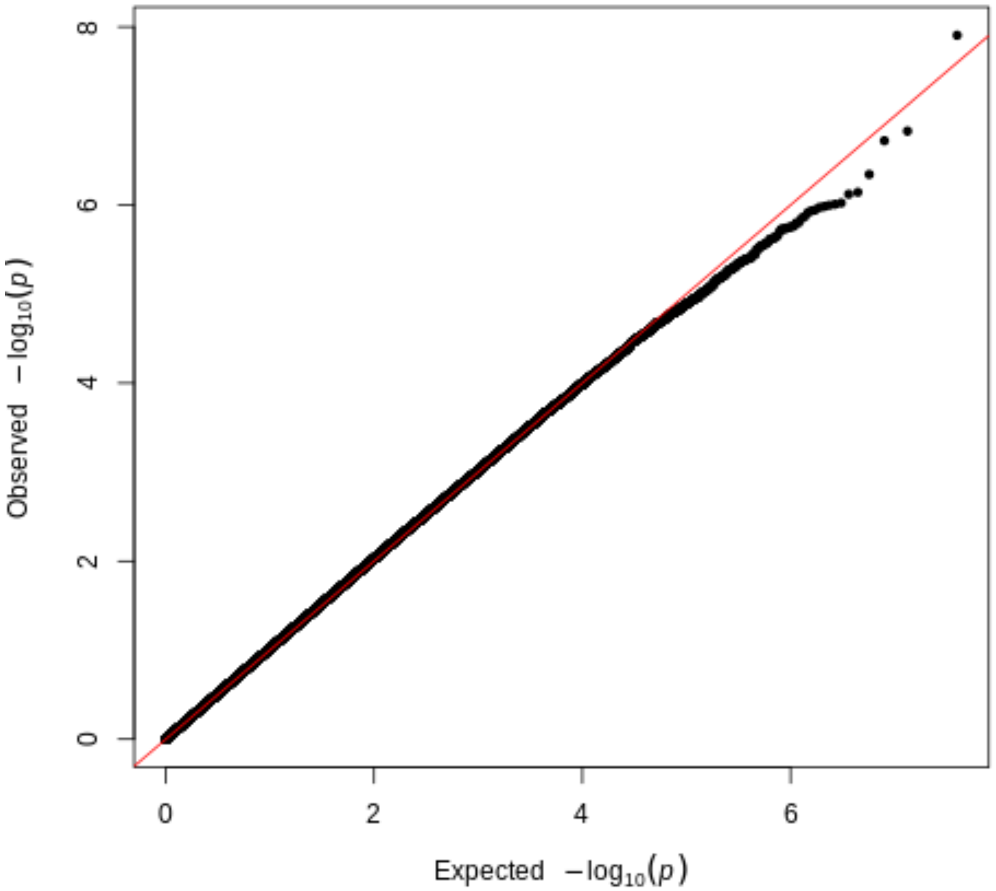


**Supplemental Figure 3.** Quantile-Quantile plot of the results of the SNP based GWAS obtained in 1,030 individuals with PDAC and 889 controls. In the Q-Q plot, we don't see genomic inflation (Lambda =1.0).

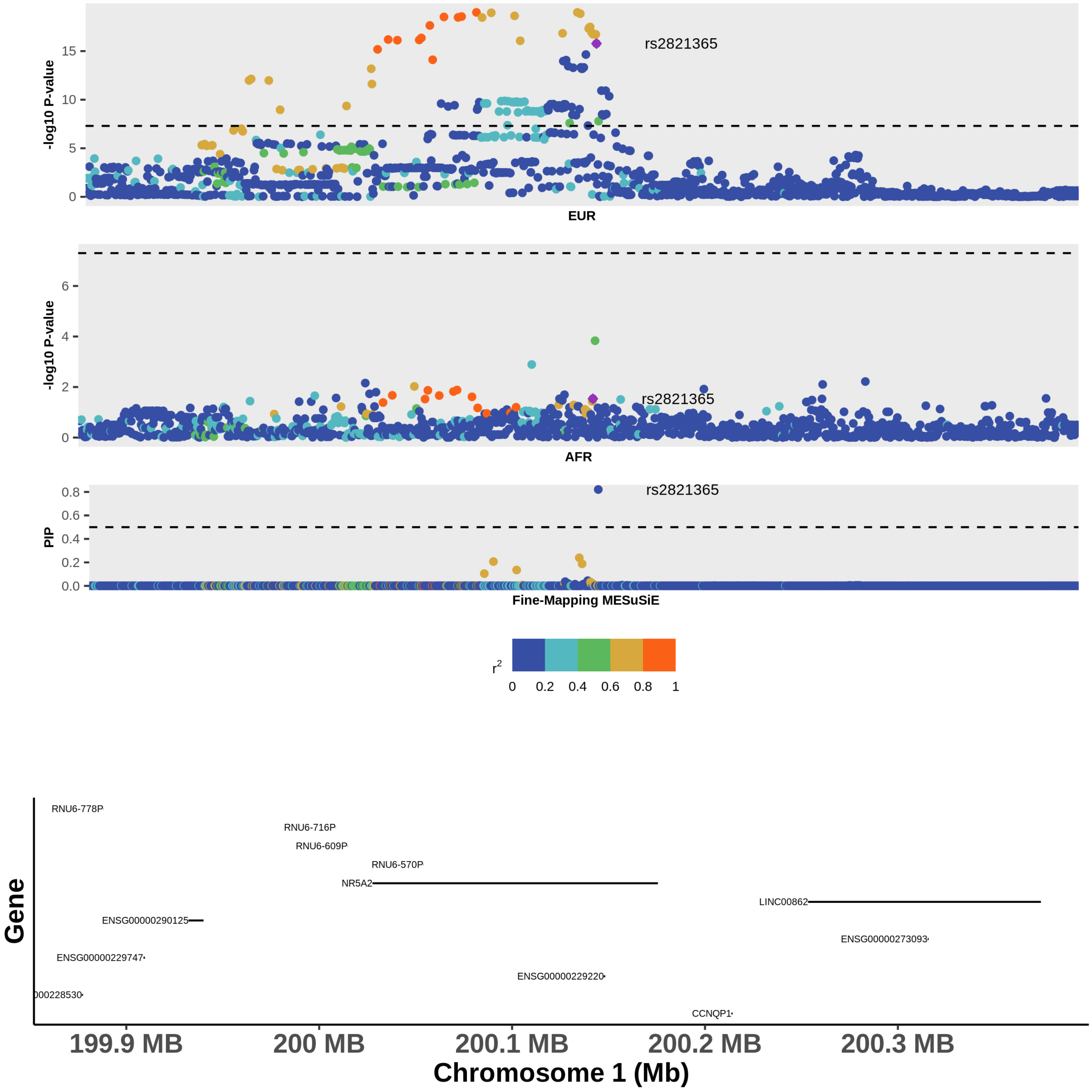


**Supplemental Figure 4**. Locus plot with the results of the fine mapping of PDAC at the 1q32.1 locus. From top to bottom, the three graphs of the upper panel display 1) the GWAS results in European (EUR) ancestry population, 2) GWAS results in African ancestry populations (AFR) and 3) results of the multi-ancestry fine mapping (Fine-Mapping MESuSiE). SNPs are colored according to their $r^2$ value with the variant with significant PIP (purple diamond) based on the color pallet annotated in the legend. Annotated SNP are those with posterior inclusion probabilities > 0.5 in the multi ancestral fine mapping. The lower panel displays gene definitions based on USCS GENCODE Genes track (version 46, May 2024) using GRCh38 coordinates.

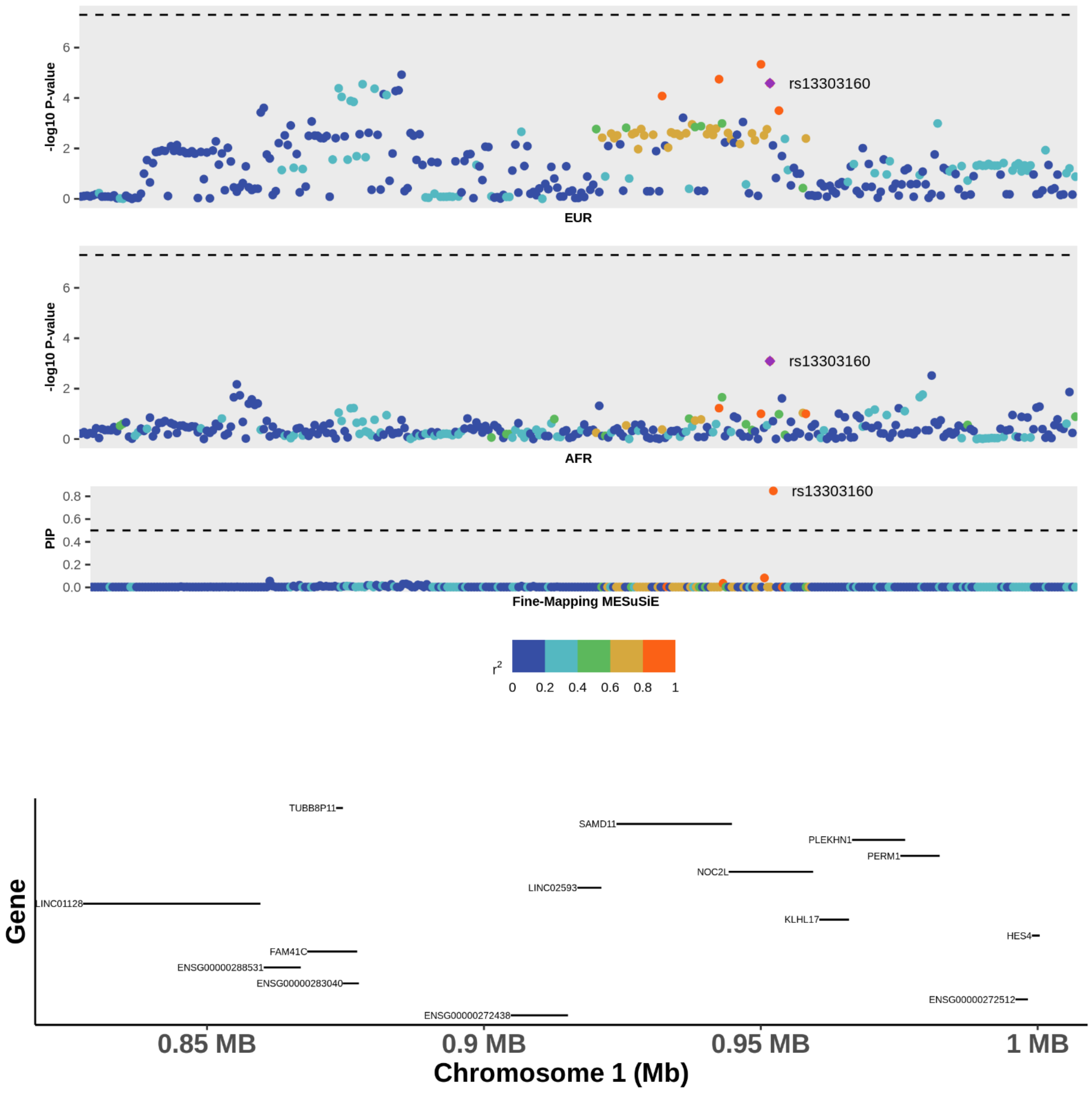


**Supplemental Figure 5**. Locus plot with the results of the fine mapping of PDAC at the 1p36.33 locus. From top to bottom, the three graphs of the upper panel display 1) the GWAS results in European (EUR) ancestry population, 2) GWAS results in African ancestry populations (AFR) and 3) results of the multi-ancestry fine mapping (Fine-Mapping MESuSiE). SNPs are colored according to their $r^2$ value with the variant with significant PIP (purple diamond) based on the color pallet annotated in the legend. Annotated SNP are those with posterior inclusion probabilities > 0.5 in the multi ancestral fine mapping. The lower panel displays gene definitions based on USCS GENCODE Genes track (version 46, May 2024) using GRCh38 coordinates.

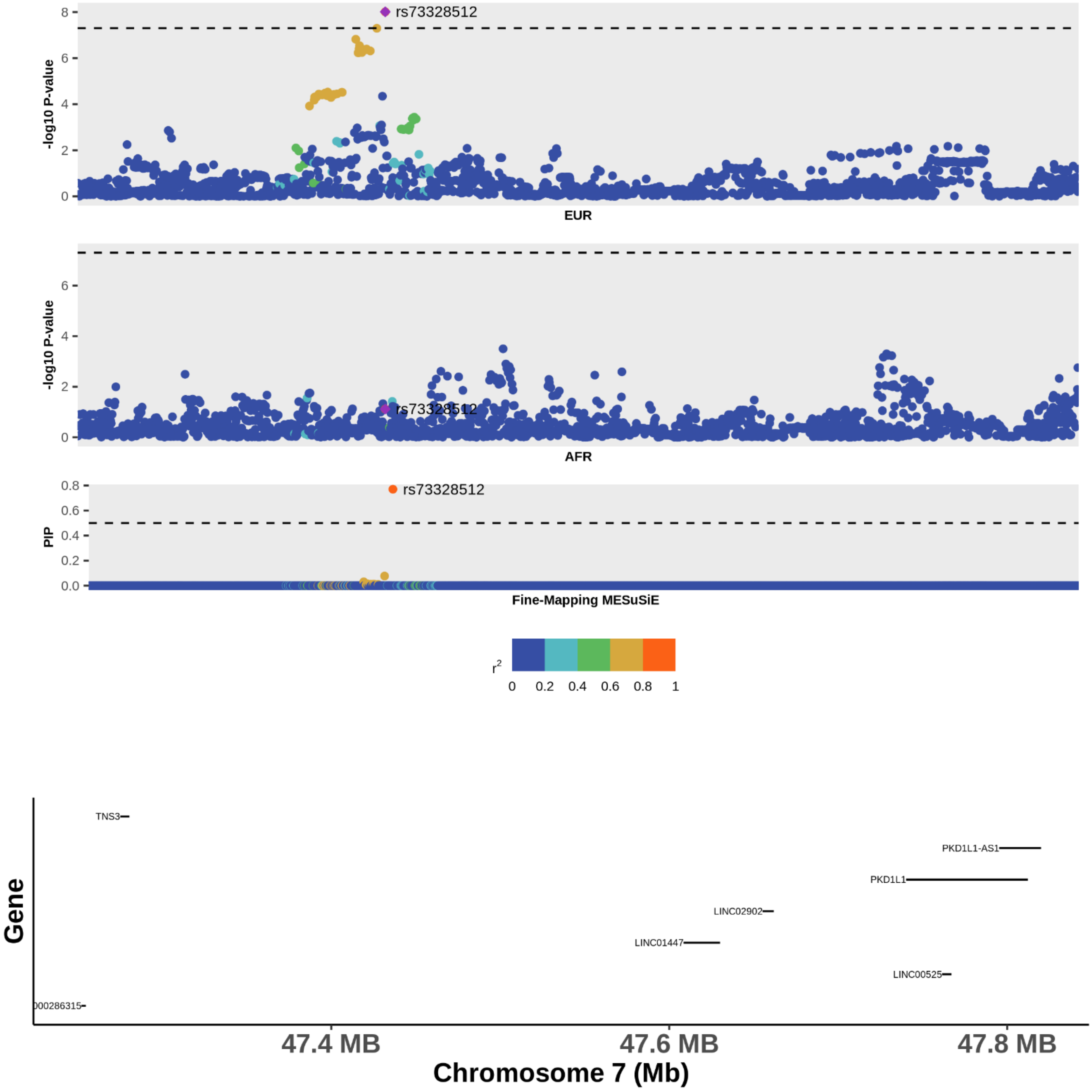


**Supplemental Figure 6**. Locus plot with the results of the fine mapping of pancreatic cancer at the 7p12.3 locus. From top to bottom, the three graphs of the upper panel display 1) the GWAS results in European (EUR) ancestry population, 2) GWAS results in African ancestry populations (AFR) and 3) results of the multi-ancestry fine mapping (Fine-Mapping MESuSiE). SNPs are colored according to their r2 value with the variant with significant PIP (purple diamond) based on the color pallet annotated in the legend. Annotated SNP are those with posterior inclusion probabilities > 0.5 in the multi ancestral fine mapping. The lower panel displays gene definitions based on USCS GENCODE Genes track (version 46, May 2024) using GRCh38 coordinates.

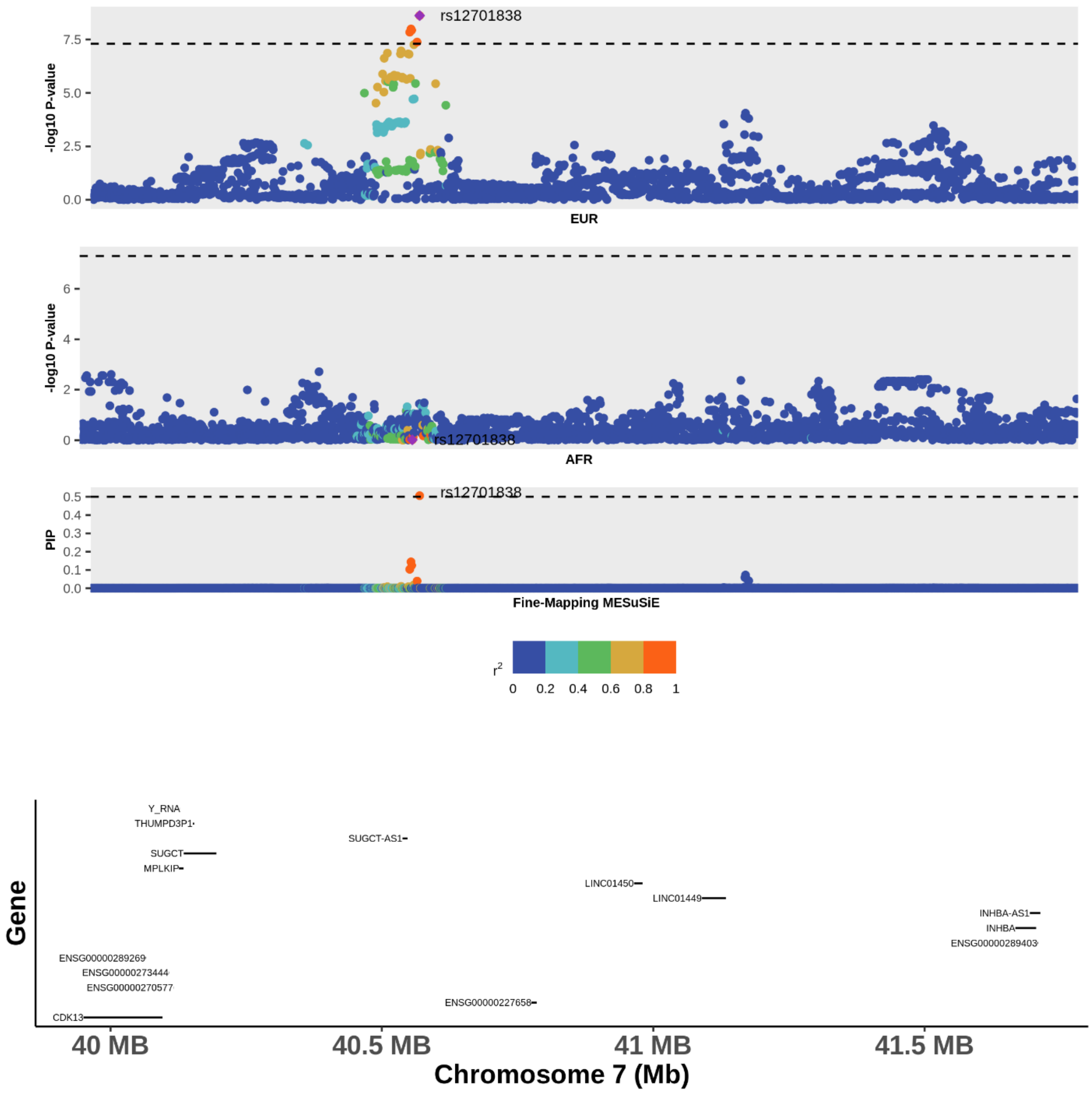


**Supplemental Figure 7**. Locus plot with the results of the multi ancestry fine mapping of pancreatic cancer at the 7p14.1 locus**.** From top to bottom, the three graphs of the upper panel display 1) the GWAS results in European (EUR) ancestry population, 2) GWAS results in African ancestry populations (AFR) and 3) results of the multi-ancestry fine mapping (Fine-Mapping MESuSiE). SNPs are colored according to their r2 value with the variant with significant PIP (purple diamond) based on the color pallete annotated in the legend. Annotated SNP are those with posterior inclusion probabilities > 0.5 in the multi ancestral fine mapping. The lower panel displays gene definitions based on USCS GENCODE Genes track (version 46, May 2024) using GRCh38 coordinates.

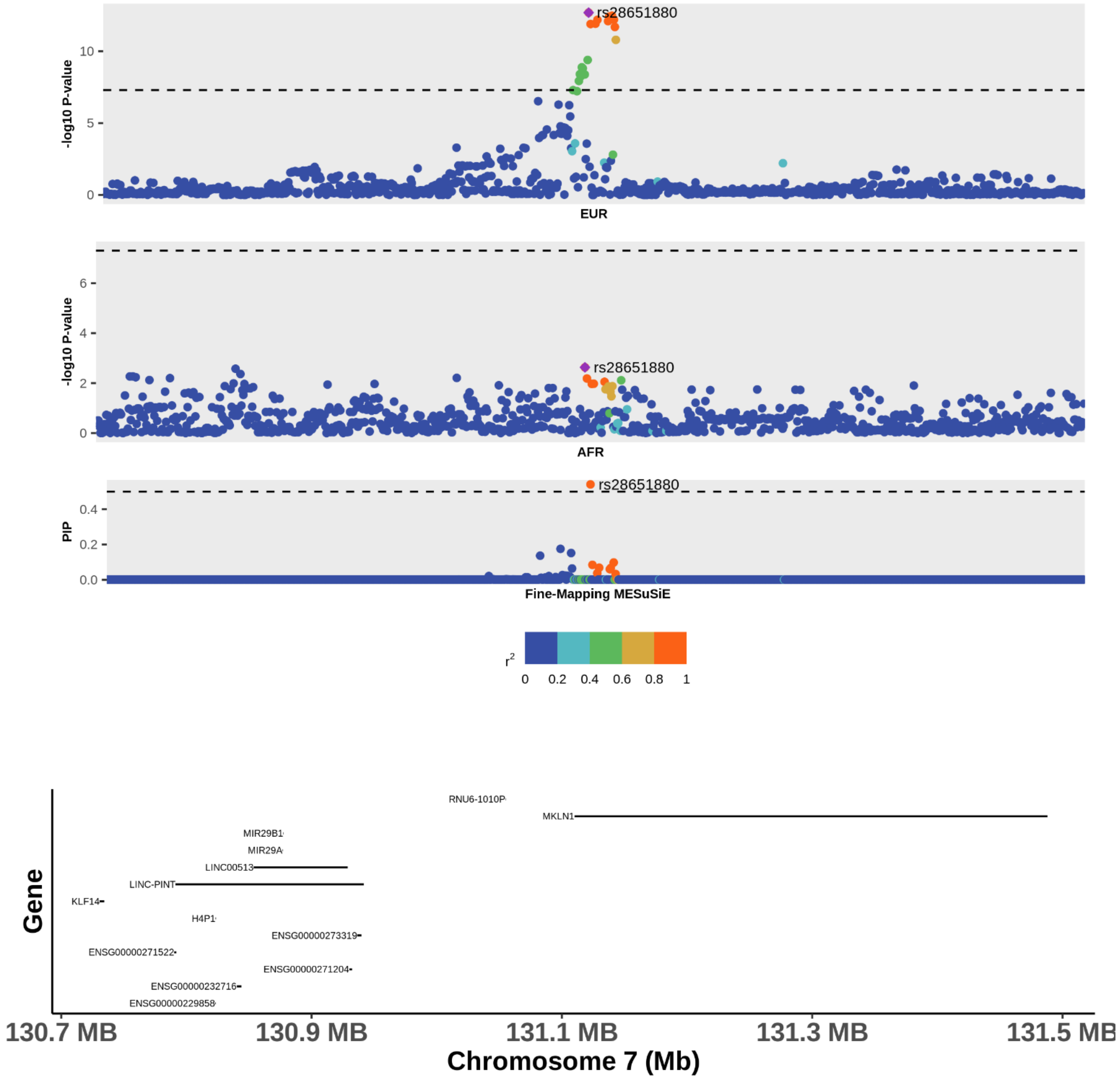


**Supplemental Figure 8**. Locus plot with the results of the fine mapping of pancreatic cancer at the 7q32.3 locus. From top to bottom, the three graphs of the upper panel display 1) the GWAS results in European (EUR) ancestry population, 2) GWAS results in African ancestry populations (AFR) and 3) results of the multi-ancestry fine mapping (Fine-Mapping MESuSiE). SNPs are colored according to their r2 value with the variant with significant PIP (purple diamond) based on the color pallete annotated in the legend. Annotated SNP are those with posterior inclusion probabilities > 0.5 in the multi ancestral fine mapping. The lower panel displays gene definitions based on USCS GENCODE Genes track (version 46, May 2024) using GRCh38 coordinates.

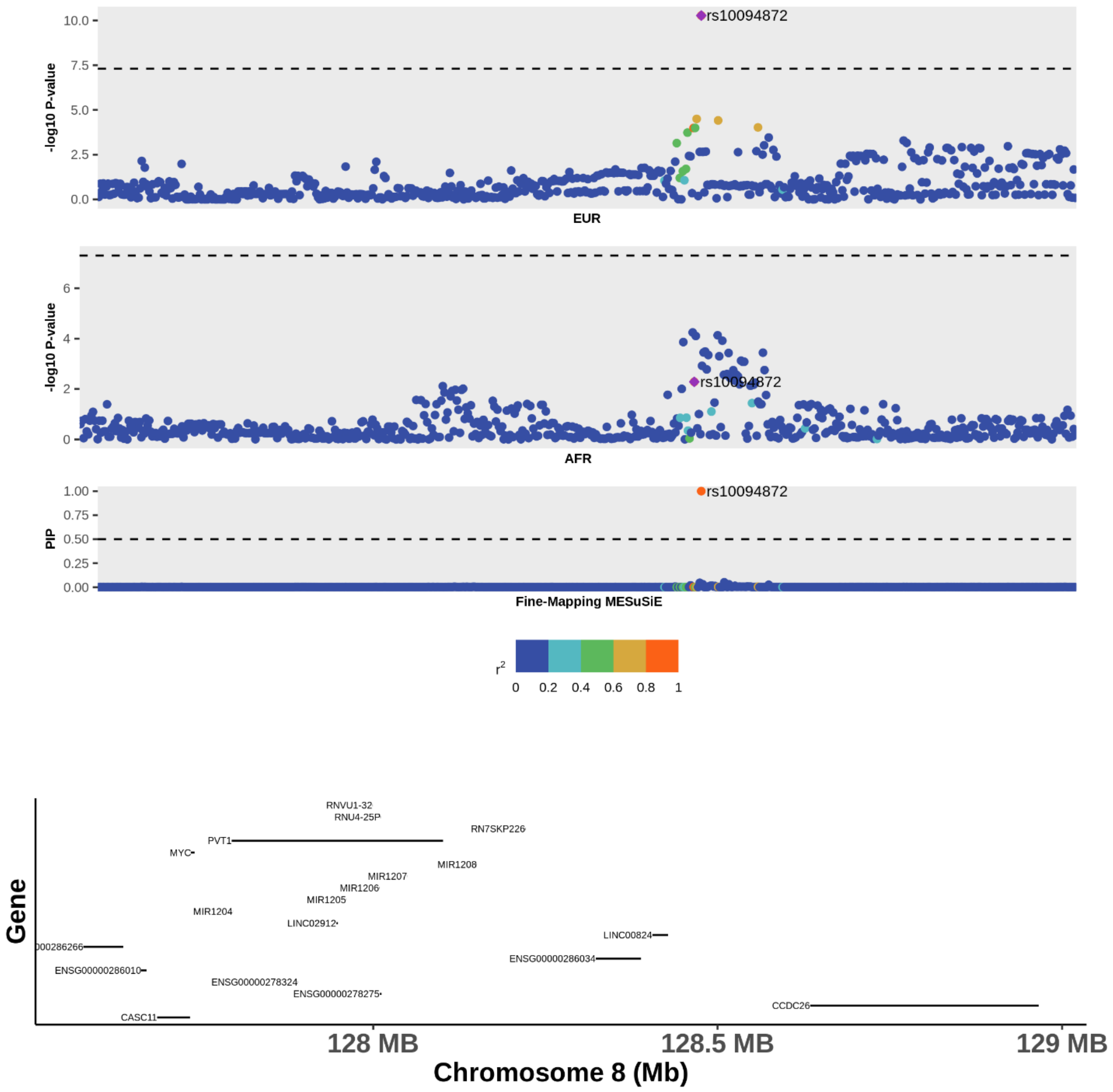


**Supplemental Figure 9**. Locus plot with the results of the fine-mapping of pancreatic cancer at the chr8q24.21 locus. From top to bottom, the three graphs of the upper panel display 1) the GWAS results in European (EUR) ancestry population, 2) GWAS results in African ancestry populations (AFR) and 3) results of the multi-ancestry fine mapping (Fine-Mapping MESuSiE). SNPs are colored according to their r2 value with the variant with significant PIP (purple diamond) based on the color pallet annotated in the legend. Annotated SNP are those with posterior inclusion probabilities > 0.5 in the multi ancestral fine mapping. The lower panel displays gene definitions based on USCS GENCODE Genes track (version 46, May 2024) using GRCh38 coordinates.

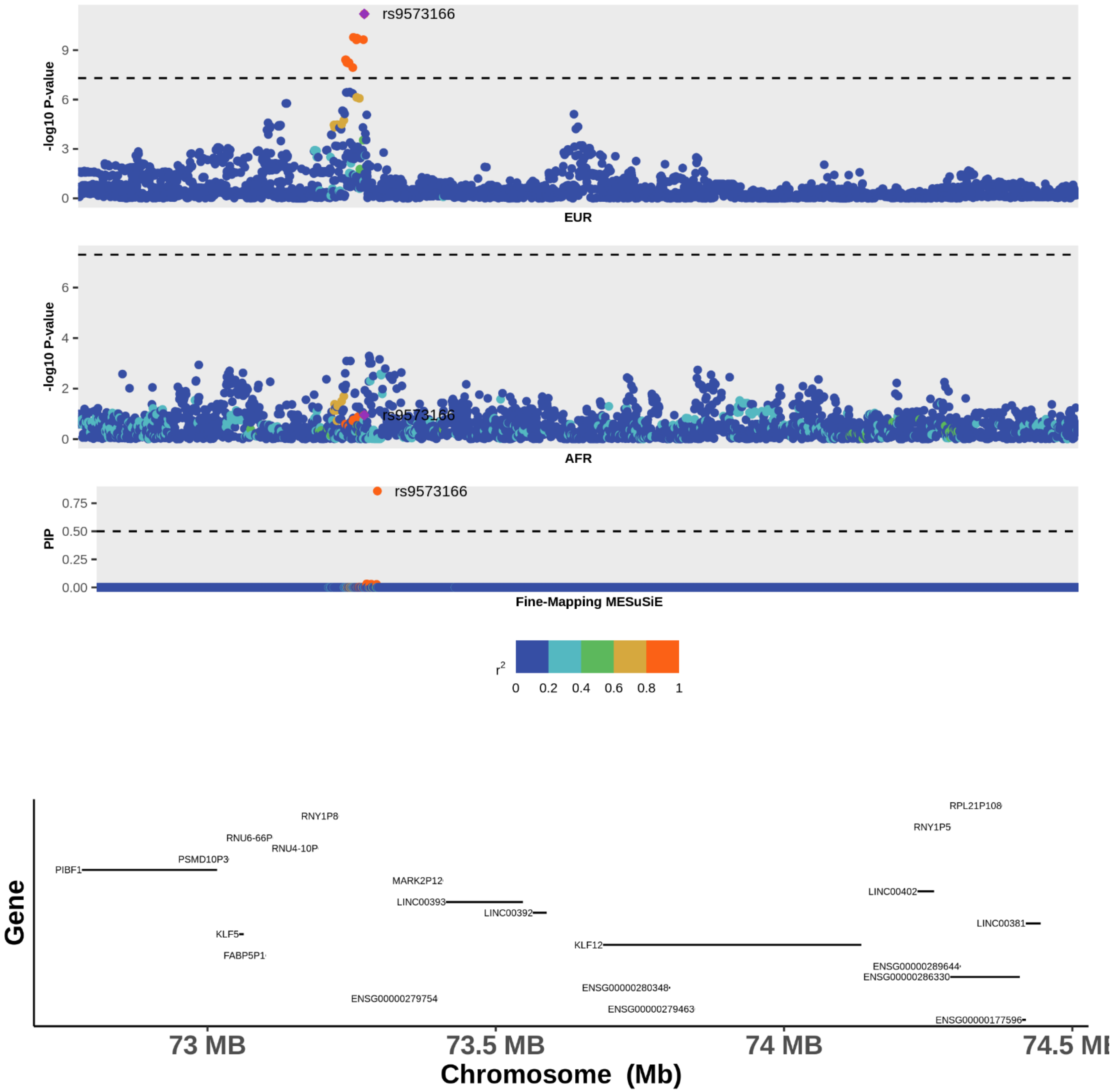


**Supplemental Figure 10**. Locus plot with the results of the fine-mapping of pancreatic cancer at the 13q22.1 locus. From top to bottom, the three graphs of the upper panel display 1) the GWAS results in European (EUR) ancestry population, 2) GWAS results in African ancestry populations (AFR) and 3) results of the multi-ancestry fine mapping (Fine-Mapping MESuSiE). SNPs are colored according to their r2 value with the variant with significant PIP (purple diamond) based on the color pallet annotated in the legend. Annotated SNP are those with posterior inclusion probabilities > 0.5 in the multi ancestral fine mapping. The lower panel displays gene definitions based on USCS GENCODE Genes track (version 46, May 2024) using GRCh38 coordinates.

**Novel Genetic Risk Loci for Pancreatic Ductal Adenocarcinoma Identified in a Genome-wide Study of African Ancestry Population**

**Supplementary Materials and Methods**

***Study Population***

Pancreatic ductal adenocarcinoma (PDAC) patients and controls belong to two large collaborative networks including sites in The Pancreatic Cancer Case Control Consortium (PANC4) and The Pancreatic Cancer Cohort Consortium. These include: American Cancer Society - Cancer Prevention Study, Columbia University, Dana Farber Cancer Institute, Fox Chase Cancer Center, Johns Hopkins Hospital, MD Anderson Cancer Center, Multiethnic Cohort Study of Diet and Health, Memorial Sloan Kettering Cancer Center, Mayo Clinic Molecular Epidemiology Case Control Study, Moffit Cancer Center, New York University Women's Health Study, Prostate, Lung, Colorectal, Ovarian Cancer Screening Trial (PLCO), University of Toronto, Sinai Health, Southern Community Cohort Study, Women's Health Initiative Yale, University of San Francisco, Give us a Clue to Cancer and Heart Disease (CLUE II). With additional controls from the INHALE study Wayne State University/Karmanos Cancer Institute [1].

***Sequencing and Genotype Quality control***

SNVs and small indels were called using GATK's 3+HaplotypeCaller joint-calling gVCF workflow. Sample replicates were checked with Picard GenotypeConcordance (https://gatk.broadinstitute.org/hc/en-us/articles/360037425091-GenotypeConcordance-Picard). All samples were found to have no less than 80% of the genome at 10x coverage and average autosomal coverage at least 20x. A multi-sample VCF of joint calls was generated with the DRAGEN Population Joint Caller (v1.2.0) [2]. Hard filters quality control removed SNVs or indels with QUAL scores less than 3. Variants were called in 1,973 experimental samples without duplicates and standard genome wide quality control was performed for the samples and markers included in the GWAS analysis [3]. Sample quality control included the exclusion of samples with sex discrepancies (n=0), unexpected duplicates (n = 6), first- and second-degree relatedness (n=6), missing age (n=4) and African ancestry proportion <.2 (n=38) with a final dataset of 1,919 individuals corresponding to 1,030 cases and 889 controls [3].

***Local and Global African Ancestry estimation***

Beagle 5.4 [4] was used to obtain phased data to be used for local ancestry estimation. Genomic position, allele order and chromosome strand of the markers in the target data was adjusted to match the reference panel using the conform-gt program (washington.edu). Reference panel used for phasing included approximately 78 million variants generated by sequencing 2,504 genomes from individuals from 16 populations worldwide included in Phase 3 sequencing of the 1000 Genomes Project [5-8]. Genetic map positions in centimorgans in the target population were estimated from GRCh38 genetic coordinates via the Beagle utility software (https://faculty.washington.edu/browning/beagle_utilities/utilities.html). Genome wide local ancestry was estimated using RFMIx [17]. This program models ancestry along the observed haplotype sequences of known or inferred ancestry. It determines both the dominant single ancestry of non-admixed individuals and decomposes the genome of admixed individuals into discrete segments originating from different ancestral populations. The ancestral populations are represented to the RFMix program [9] using a reference panel of present-day populations as proxies for populations which contributed to today's admixed populations. Utah residents with Northern and Western European ancestry (CEU) and samples from Yoruba in Ibadan, Nigeria (YRI) included in the 1000 Genomes Project were used as reference populations for European and African descent, respectively [5]. Only unrelated individuals were retained, resulting in 98 CEU and 97 YRI unrelated reference samples [10]. GRCh38 genetic linkage information and genetic map was used by RFMix to define windows for local ancestry assignment. The number of alleles inherited from African and European ancestral populations for each individual was inferred using a window size of 0.2 cM assuming 6 generations since the event of admixture reflecting estimates from prior studies in the United States [11]. Global admixture proportions were estimated for each individual genome-wide average local ancestry. Global estimates and local ancestry proportions were summarized and graphed using customized scripts in R (19).

Global inferred African ancestry was estimated to account for population structure in downstream analyses. Across the study population, estimated African ancestry proportions ranged from 0.24 to 0.82 (**Supplemental Figure 2**). The proportion of African ancestry did not differ (P value =0.25) between individuals with PDAC (Median 0.679; IQR= 0.602, IQR-0.727 controls **(**Median=0.675; IQR= 0.609-0.725), indicating similar ancestry proportions in both groups **(Table 1, Supplementary Figure 2)**. Sensitivity analyses restricting to studies with >20 cases and >20 controls showed similar distributions, indicating no systematic differences that could confound local ancestry analyses (P value = 0.82).

***<u>Association of Local Ancestry and PDAC (Admixture Mapping).</u>***

For the association analysis, we estimated principal components and a genetic relatedness matrix (GRM) using 113,467 autosomal markers with MAF > 0.40 selected as unlinked using standard parameters (sliding window of 500 SNPs, advanced in steps of 5 SNPs, with a pairwise $r^2$ threshold of 0.2) in PLINK 1.9 [12] ) using the PC-AiR program [14].PC-AiR performs a principal components Analysis on genome-wide SNP data for the detection of population structure in a sample that may contain known or cryptic relatedness. Unlike standard PCA, PC-AiR accounts for cryptic or family relatedness in the sample to provide accurate ancestry inference that is not confounded by family structure. PC-AiR uses measures of ancestry divergence estimated using KING-Robust algorithm [13] to partition samples into related and unrelated ancestry representative sets. Estimation of pairwise kinship coefficients adjusted for population structure were obtained using PC-Relate [14]. PC-Relate uses ancestry representative principal components to adjust for population structure/ancestry and accurately estimate measures of recent genetic relatedness such as kinship coefficients, IBD sharing probabilities, and inbreeding coefficients [14]. We obtained principal components that are robust to relatedness and ancestry as well as an adjusted genetic relationship matrix (GRM) by estimating PCs starting with the original GRM obtained in the first round of analysis. Using this iterative procedure, we obtained kinship coefficients adjusted for population structure and admixture, which were used in the logistic mixed model of admixture mapping.

#### ***Genome Wide Association Analysis***

Genome Wide Association Analysis was performed using SAIGE (Scalable and Accurate Implementation of GEneralized mixed model) [15]. This program uses two steps: (1) fitting the null logistic mixed model to estimate the variance component and other parameters; (2) testing for the association between each genetic variant and PDAC by applying saddle point approximation to the score test statistics. Step 1 iteratively estimated the model parameters average information restricted maximum likelihood (AI-REML) algorithm. In the null model, we included the genetic relationship matrix (GRM) based on the raw genotypes, age in 5-year age categories (as described in **Table 1**), sex and one principal component. After fitting the null logistic mixed model, we obtained estimates of the random effects for each individual. The ratio of the variances of the score statistics with and without incorporating the variance components for the random effects is calculated using a subset of randomly selected genetic variants. In step 2, for each variant, the variance ratio is used to calibrate the score statistic variance that does not incorporate variance components for random effects. SAIGE next approximates the score test statistics using the saddle point approximation to obtain more accurate P values than the normal distribution.

***Multi-Ancestry Fine Mapping***

For fine mapping we used the multi-ancestry sum of the single effects model (MESuSiE) program [16]. It assumes the presence of both shared putative causal SNPs that display non-zero effects in both ancestries and ancestry-specific causal SNPs that display non-zero effects only in one ancestry [18].This probabilistic multi-ancestry fine-mapping method builds upon the sum of single effect model and extends the normal assumption on the effect size of causal SNP to multivariate normal, capturing correlation across ancestries. It uses centered genotype matrices and the sum of the single effect models in each population to determine the putative causal variant for a standardized phenotype. This probabilistic multi-ancestry fine-mapping method has improved accuracy and resolution of fine-mapping by leveraging association information across ancestries as cross-population fine-mapping has the potential to improve power and resolution by capitalizing on the genomic diversity across ancestries. MESuSiE uses summary statistics as input in the form of SNP information, marginal effect size, standard error, Z-scores, and sample size. It accounts for the diverse linkage disequilibrium pattern observed in different ancestries based on an ancestry-specific linkage disequilibrium matrix and explicitly models both shared and ancestry-specific causal SNPs.

***Genotyping, quality control (QC) and association analysis for GWAS in individuals of European genetic similarity***

Genotyping for individuals included in the PanScan Study (https://dceg.cancer.gov/research/cancer-types/pancreas/panscan) was performed at the Cancer Genomics Research Laboratory (CGR) of the National Cancer Institute (NCI) of the National Institutes of Health (NIH) using different genotyping platforms as described in detail in reference [17]. In brief, the Illumina HumanHap series arrays (Illumina HumanHap550 Infinium II, Human 610-Quad) were used for PanScan I [18] and II [19], respectively, and the Illumina Omni series arrays (OmniExpress, Omni1M and Omni2.5) for PanScan III[20] as previously described [17]. Due to the large overlap of variants on genotyping arrays for PanScan I and II, these two datasets were analyzed together. The PanScan III GWAS dataset was analyzed separately. Genotyping for the PanC4 GWAS was performed at the Johns Hopkins Center for Inherited Disease Research (CIDR) using the Illumina HumanOmniExpressExome-8v1 array and was imputed separately. As Genotype QC prior to imputation: variants with one or more multi-character allele codes or with single-character allele codes outside of {'A', 'C', 'G', 'T', 'a', 'c', 'g', 't', <missing code>} were excluded, and duplicate variants were merged using PLINK [21]; variants on the respective arrays above were remapped, and strands were flipped if necessary using Will Rayner's strand files (https://www.chg.ox.ac.uk/~wrayner/strand/); a two-stage filtering process was applied, using a completion rate threshold of 0.80 for both samples and loci, followed by a 0.95 threshold for both; samples with discordant gender (self-reported gender vs. genetic gender) were excluded; variants with minor allele frequency (MAF) < 0.01 or Hardy Weinberg Equilibrium (HWE) $P < 1x10^{-6}$ for control individuals or $P < 1x10^{-12}$ for cases were excluded; variants with allele frequency differences > 0.2 between our data and the TOPMed reference panel, those not in the TOPMed reference panel, or A/T and G/C variants on ambiguous DNA strand (MAF > 0.4) were excluded (McCarthy Group Tools: https://www.chg.ox.ac.uk/~wrayner/tools/). Principal component analysis (PCA) was performed using PLINK [21] and the top 20 PCs remained for further analysis. A logistic regression model using the glm (generalized linear model) R package was used to test if the PCs were significantly associated with the PDAC phenotype; only significant PCs were used in the follow up association analysis. After genotype quality control, genotypes were imputed using the TOPMed imputation reference panel [22] (version R2 on human genome build GRCh38) via the Michigan Imputation Server [23-25]. As Post-imputation genotype QC, ancestry was assessed using the Genotyping Library and Utilities (GLU- http://code.google.com/p/glu-genetics/) struct.admix module based on the method by Pritchard et al. [26] and samples with < 80% European ancestry were excluded; variants with MAF < 0.001, imputation quality score ($R^2$) <

0.3 were excluded; samples with completion rates < 90% or heterozygosity rates +/- 3 standard deviation from the mean were excluded; samples that represented ≤ 2nd degree relatives (within or across PanScan I+II and PanScan III GWAS phases) were excluded (the sample with the highest completion rate was retained); duplicate samples in the PanScan I+II and PanScan III GWAS phases were excluded from PanScan III. For PanScan III, optmatch [27]was used to select matched controls for cases (1:3 case-to-control ratio) based on genetic principal components. Association analysis for each GWAS phase was performed using logistic regression in SNPTEST(v2.5.4-beta3) [28] based on the imputed genetic allele dosages and the "Frequentist" (additive) approach, with adjustment for the following covariates: PanScan I+II: age, gender, significant principal component(s), study; 2) PanScan III: age, gender, significant principal component(s), and geographic region. Age was categorized into the following age groups: < 51, 51-60, 61-70, 71-80 or >80). Geographic region for PanScan III was defined based on the geographic location of participating studies as follows: REGION_US (United States): AgHealth, CPS-II, DFCI, HPFS, MEC, NHS, NYU-WHS, PHS, PLCO, SELECT, VITAL, WHI; REGION_CNE (central and northern Europe): ATBC, EPIC, PANDoRA-Heidelberg, MCCS (Melbourne); and REGION_SE (southern Europe): SBCS (Spain controls), PANKRAS-II (cases) as per the original PanScan III GWAS analysis. 3) PANC4 was adjusted for, age, sex, and significant principal components. Genotype uncertainty was processed using a missing data likelihood score test by SNPTEST(-method score). Meta-analysis using the fixed-effects inverse-variance method based on $\beta$ estimates and SEs (http://csg.sph.umich.edu/abecasis/metal/) was conducted.

**Supplemental acknowledgements**

This work was supported by U01CA247283 and X01HG010459 and federal funds from the National Cancer Institute (NCI), US National Institutes of Health (NIH) under contract number HHSN261200800001E.

This project has been funded in whole or in part with Federal funds from the National Cancer Institute, National Institutes of Health, under NCI Contract No. 75N910D00024. The content of this publication does not necessarily reflect the views or policies of the Department of Health and Human Services, nor does mention of trade names, commercial products, or organizations imply endorsement by the U.S. Government.

The authors acknowledge the research contributions of the Cancer Genomics Research Laboratory for their expertise, execution, and support of this research in the areas of project planning, wet laboratory processing of specimens, and bioinformatics analysis of generated data.

The work at Johns Hopkins University was supported by the NCI Grants P50CA062924 and R01CA154823. Additional support was provided by the Lustgarten Foundation, Susan Wojcicki and Dennis Troper and the Sol Goldman Pancreas Cancer Research Center.

The WHI program is funded by the National Heart, Lung, and Blood Institute, National Institutes of Health, U.S. Department of Health and Human Services through 75N92021D00001, 75N92021D00002, 75N92021D00003, 75N92021D00004, 75N92021D00005." The authors thank the WHI investigators and staff for their dedication, and the study participants for making the program possible. A full listing of WHI investigators can be found at: https://www-whi-org.s3.us-west-2.amazonaws.com/wp-content/uploads/WHI-Investigator-Long-List.pdf

Data collection at the University of Toronto was supported by the CIHR Canada Research Chair and from the Canadian Cancer Society Research Institute.

The MD Anderson case control study was supported by NIH R01 CA98380 and R01 CA181244

The Southern Community Cohort Study is supported by a grant from NIH (U01 CA202979), the Shanghai Women's Health Study and Shanghai Men's Health Study were supported by NIH grants UM1CA182910 and UM1CA173640.

The Yale (CT) pancreas cancer study is supported by National Cancer Institute at the U.S.National Institutes of Health, grant 5R01CA098870. The cooperation of 30 Connecticut hospitals, including Stamford Hospital, in allowing patient access, is gratefully acknowledged. The Connecticut Pancreas Cancer Study was approved by the State of Connecticut Department of Public Health Human Investigation Committee. Certain data used in that study were obtained from the Connecticut Tumor Registry in the Connecticut Department of Public Health. The authors assume full responsibility for analyses and interpretation of these data

The NYU-WHI study was funded by NIH R01 CA098661, UM1 CA182934 and center grants P30 CA016087 and P30 ES000260.

The authors express sincere appreciation to all Cancer Prevention Study-II participants, and to each member of the study and biospecimen management group. The authors would like to acknowledge the contribution to this study from central cancer registries supported through the Centers for Disease Control and Prevention's National Program of Cancer Registries and cancer registries supported by the National Cancer Institute's Surveillance Epidemiology and End Results Program.

The ATBC Study was supported by the Intramural Research Program, Division of Cancer Epidemiology and Genetics of the U.S. National Cancer Institute (NCI), National Institutes of Health.

Cancer incidence data for CLUE were provided by the Maryland Cancer Registry, Center for Cancer Surveillance and Control, Department of Health and Mental Hygiene, 201 W. Preston Street, Room 400, Baltimore, MD 21201, http://phpa.dhmh.maryland.gov/cancer, 410-767-4055. We acknowledge the State of Maryland, the Maryland Cigarette Restitution Fund, and the National Program of Cancer Registries of the Centers for Disease Control and Prevention for the funds that support the collection and availability of the cancer registry data." We thank all the CLUE participants.

The IARC/Central Europe study was supported by a grant from the US National Cancer Institute at the National Institutes of Health (R03 CA123546-02) and grants from the Ministry of Health of the Czech Republic (NR 9029-4/2006, NR9422-3, NR9998-3, MH CZ-DRO-MMCI 00209805).

The Queensland Pancreatic Cancer Study was supported by a grant from the National Health and Medical Research Council of Australia (NHMRC) (Grant number 442302). RE Neale is supported by a NHMRC Senior Research Fellowship (#1060183).

The Yale (CT) pancreas cancer study is supported by National Cancer Institute at the U.S. National Institutes of Health, grant 5R01CA098870. The cooperation of 30 Connecticut hospitals, including Stamford Hospital, in allowing patient access, is gratefully acknowledged. The Connecticut Pancreas Cancer Study was approved by the State of Connecticut Department of Public Health Human Investigation Committee. Certain data used in that study were obtained from the Connecticut Tumor Registry in the Connecticut Department of Public Health. The authors assume full responsibility for analyses and interpretation of these data.

Assistance with genotype data quality control was provided by Cecelia Laurie and Cathy Laurie at University of Washington Genetic Analysis Center

The American Cancer Society (ACS) funds the creation, maintenance, and updating of the Cancer Prevention Study II cohort.

Cancer incidence data for CLUE were provided by the Maryland Cancer Registry, Center for Cancer Surveillance and Control, Department of Health and Mental Hygiene, 201 W. Preston Street, Room 400, Baltimore, MD 21201, http://phpa.dhmh.maryland.gov/cancer, 410-767-4055.

We acknowledge the State of Maryland, the Maryland Cigarette Restitution Fund, and the National Program of Cancer Registries of the Centers for Disease Control and Prevention for the funds that support the collection and availability of the cancer registry data.” We thank all the CLUE participants.

Melbourne Collaborative Cohort Study (MCCS) cohort recruitment was funded by VicHealth and Cancer Council Victoria. The MCCS was further augmented by Australian National Health and Medical Research Council grants 209057, 396414 and 1074383 and by infrastructure provided by Cancer Council Victoria. Cases and their vital status were ascertained through the Victorian Cancer Registry and the Australian Institute of Health and Welfare, including the National Death Index and the Australian Cancer Database.

The NYU study (AZJ and AAA) was funded by NIH R01 CA098661, UM1 CA182934 and center grants P30 CA016087 and P30 ES000260.

The Physicians' Health Study was supported by research grants CA-097193, CA-34944, CA-40360, HL-26490, and HL-34595 from the National Institutes of Health, Bethesda, MD USA.

Health Professionals Follow-up Study is supported by NIH grant UM1 CA167552. from the National Cancer Institute, Bethesda, MD USA

Nurses’ Health Study is supported by NIH grants UM1 CA186107, P01 CA87969, and R01 CA49449 from the National Cancer Institute, Bethesda, MD USA

Additional support from the Hale Center for Pancreatic Cancer Research, U01 CA21017 from the National Cancer Institute, Bethesda, MD USA , and the United States Department of Defense CA130288, Lustgarten Foundation, Pancreatic Cancer Action Network, Noble Effort Fund, Peter R. Leavitt Family Fund, Wexler Family Fund, and Promises for Purple to B.M. Wolpin.

SELECT study is supported by National Institutes of Health grant award number U10 CA37429 (CD Blanke), and UM1 CA182883 (CM Tangen/IM Thompson). The authors thank the site investigators and staff and, most importantly, the participants from PCPT and SELECT who donated their time to this trial.

This study utilized the high-performance computational capabilities of the Biowulf Linux cluster at the NIH, Bethesda, MD, USA (http://biowulf.nih.gov).
